# Developing Buoyant-Analyte-Magnetic (BAM) Assays for Ultrasensitive Yet Rapid Point-of-Care Detection

**DOI:** 10.64898/2026.06.15.26355555

**Authors:** Chuanlei Wang, Emory Satterfield, Nicholas Erwin, Jose Correa, Wesley Wampler, Phillip Moschella, Delphine Dean, Jeffrey N. Anker

## Abstract

Rapidly detecting infectious diseases such as Covid-19 is essential to control outbreaks and treat patients early. However, no available screening method combines low cost, portability, speed (<20 min, ideally <5 min), and ultrasensitivity (e.g., <1 virus/µL): lateral flow assays are fast, portable, and inexpensive but insensitive, whereas ultrasensitive assays require centralized labs with long turnaround times. We recently developed an ultrasensitive immunoassay that captures, separates, and counts saliva biomarker molecules using buoyant microbubbles and magnetic microspheres, but the original assay took 55 minutes and was not readily deployable. Here, we redesigned the assay protocol and reader for emergency medicine and mobile care by streamlining the workflow, collecting saliva with larger swabs, filtering it through a 10 µm cap, and using larger microbubbles to accelerate flotation. A paramedic successfully ran the assay on the back of a parked medical van in 3.5 minutes (spit-to-results) while achieving a 1.3 fg/mL analytical detection limit for SARS-CoV-2 nucleocapsid protein (∼0.04 virus1/µL). The assay remained positive across 9 orders of magnitude. We describe the challenges and opportunities ahead for point-of-care deployment.

## 1. Main

Diagnosing communicable diseases, such as Covid-19, rapidly and at early stages of infection is essential to effectively treat patients and contain outbreaks. However, early-stage diagnosis remains challenging because disease biomarkers are often present at trace concentrations.^1–3^ Inexpensive, user-friendly tests, such as lateral flow assays (LFA), can be run rapidly at the point-of-care (POC) but lack sensitivity at early stages.^2,4^ Conversely, centralized labs can run ultrasensitive immunoassays and polymerase chain reaction (PCR) assays, but instrument size, protocol complexity, protocol speed, and instrument & reaction costs make them unsuitable for POC detection.

The tradeoff between sensitivity and POC use is highlighted in the World Health Organization (WHO) 2020 target product profile (TPP) for Covid-19 diagnostics,^5^ which defines separate performance goals for laboratory-based diagnostics and rapid screening. The minimum acceptable limit of detection (LOD) for diagnostic assays was 1 cp/µL with a 4-hour test, with a desirable goal of 0.1 cp/µL within 45 minutes, whereas rapid screening assays targeted <10³ RNA cp/µL within 40 minutes, with a desirable target of <10 cp/µL within 20 minutes. Although not envisioned in the TPP, an inexpensive <5-minute assay would be especially useful for population screening during outbreak containment. Quantitatively, commercial LFAs have LODs equivalent to a range of 2×10^2^ to 6×10^3^ cp/µL.^2^ Some of these meet the WHO’s acceptable POC specifications, although not the desirable target. Clinically, LFAs have ∼70% sensitivity vs. gold-standard RT-qPCR in symptomatic patients (∼50% for asymptomatic patients).^4^ Compared to ultra-sensitive lab-based assays, LFAs are typically 2 to 4 orders of magnitude less sensitive.^2,3,6^ While LFAs can identify highly infectious individuals in community settings, their limited analytical sensitivity reduces detection during early stages of infection and potentially enables transmission, especially to people in prolonged proximity (e.g., family transmission)^2,7^ and to susceptible immunodeficient individuals.^8,9^

Ultrasensitive diagnostic assays are typically confined to centralized laboratories, causing delays for specimen transportation and batch processing, with results reported hours to days after patients provide specimens.^10^ These laboratory-based methods include high-performance protein immunoassays such as the Quanterix single-molecule array (Simoa)^11,12^ and nucleic acid amplification tests (NAATs), particularly RT-qPCR, which is the gold standard for Covid-19 diagnosis.^4,13^ To bridge the sensitivity/POC gap, portable isothermal NAATs, such as loop-mediated isothermal amplification (LAMP) and nicking enzyme amplification reaction (NEAR), have been developed. However, compared to LFAs they are relatively slow (typically >30 min), often require refrigerated storage, and remain relatively expensive due to the cost of the enzymes and readout equipment.^14^ They also typically exhibit higher LODs than lab-based techniques partly due to artifacts such as primer dimerization, (∼10–100 RNA cp/µL),^15^ meeting the WHO’s POC target but not the diagnostic target. Thus, combining diagnostic sensitivity with low-cost POC deployment remains challenging in real-world infectious scenarios.

Figure 1 compares common assays based on ultrasensitive vs. POC needs, and whether they can detect antigens (usually cheaper and required for some applications). Our BAM assay combines ultrasensitive diagnostics with a user-friendly, rapid (<5-minute) antigen test. During the procedure, analyte molecules in saliva (in this case, SARS-CoV-2 nucleocapsid protein, N-protein) bind to antibodies on both buoyant microbubbles and magnetic microspheres, forming buoyant-analyte-magnetic (BAM) sandwich complexes. These complexes are separated from unbound buoyant and magnetic labels in solution using a combination of buoyant and magnetic forces (Figure 2). A flashlight illuminates the BAM complexes, which scatter light intensely, and a simple digital camera counts them to detect and quantify N-protein in the specimen. Recently, we showed that BAM assays could detect trace N-protein levels in patient saliva, but our protocol had too many steps and took too long (55 minutes) for POC testing.^16^ Here, we improved the assay for POC use by making the platform portable, increasing the microbubble size, simplifying the workflow, and reducing assay time to 3.5 minutes. We also showed that the assay was simple enough for emergency medicine technicians and paramedics to run in Emergency Department (ED) labs and on the trunk of a parked emergency response vehicle.

**Figure 1:**
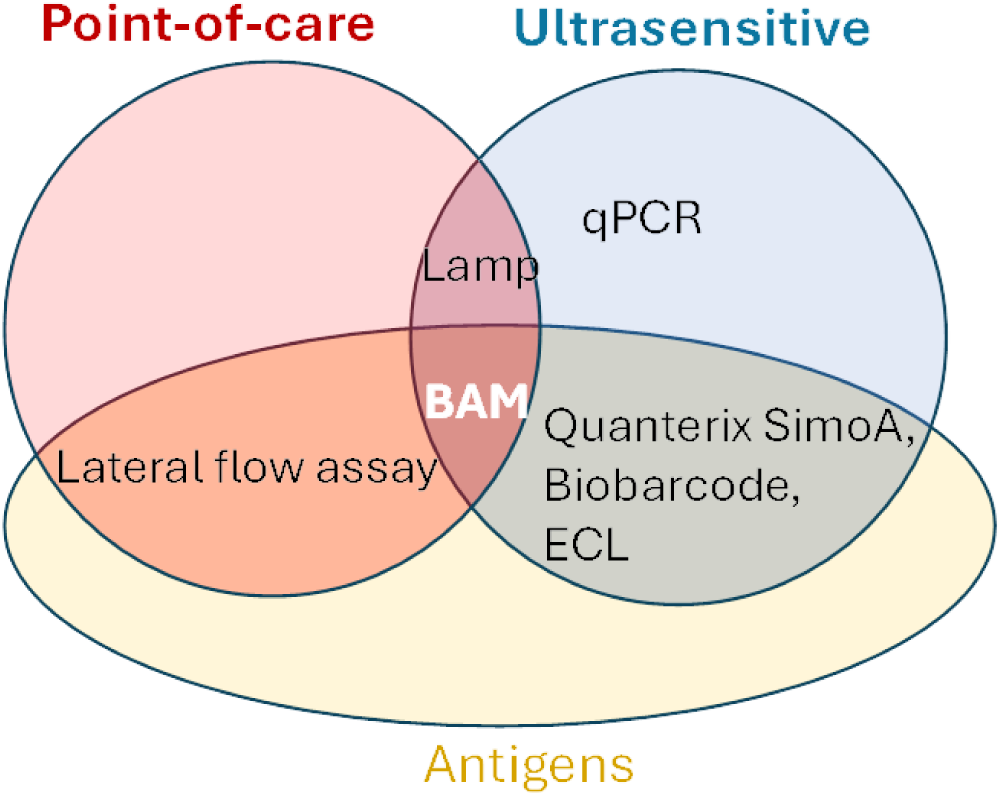
Venn Diagram categorizing BAM assays as ultrasensitive, antigen detecting at the point-of-care. Lamp: Loop-mediated isothermal amplification; Simoa: single molecule array; ECL: electrochemiluminescence; ELISA: Enzyme-linked immunosorbent assay.

**Figure 2:**
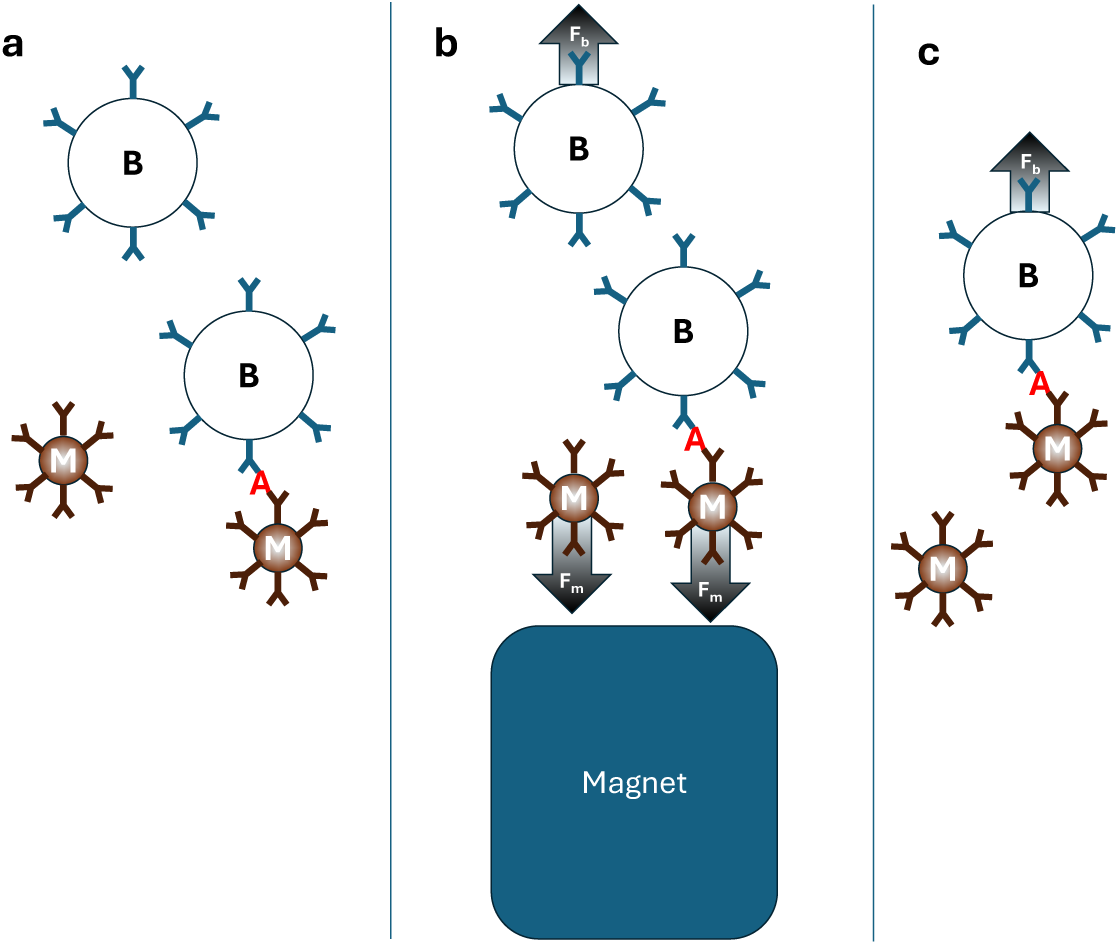
BAM assay steps schematic (not to scale). **a)** Buoyant microbubbles (labeled B), analyte (A), and magnetic microspheres (M) are mixed and form BAM complexes. The “Y” shapes represent antibodies. **b)** A magnet pulls down the BAM complexes and unbound magnetic microspheres, while unbound microbubbles float away. **c)** Removing the magnet releases the BAM complexes, which float upwards and are counted. Note, the Methods Section provides a fuller protocol for saliva analysis at the POC, and Supplementary Figure 9 is a more detailed instruction booklet.

## 2. Results

Previously, our proof-of-principle BAM assay achieved the best-reported LOD in 5 µL patient saliva (0.58 fg/mL).^16^ However, this assay used relatively complex instrumentation and multiple steps, with a 55-minute spit-to-analysis time. Here, we simplified the assay, improved instrument portability, and reduced turnaround time to 3.5 minutes. To achieve these goals, we iterated the design and protocol twice (see Table 1). We then evaluated assay reproducibility by an Emergency Department technician and a paramedic in POC environments.

**Table 1:** Comparison of the original assay and two iterations. Yellow corresponds to the original assay specification. Orange is an intermediate specification. Green is the final specification.

|  |  | Assay Iteration |  |  |
| --- | --- | --- | --- | --- |
|  |  | Original Lab Assay<br>(from ref <sup>16</sup>). | Iteration 1 (large<br>microbubbles) | Iteration 2 (large<br>microbubbles +<br>improved instrument<br>and POC protocol) |
| Disposables | Parameter / Feature |  |  |  |
| | Buoyant Bead size | 15 $\mu\text{m}$ | 27 $\mu\text{m}$ | 27 $\mu\text{m}$ |
|  | Antibody<br>functionalization/BB | ~40,000/Bead | ~800,000/Bead | ~800,000/Bead |
|  | Number of BB in incubation | ~750,000 | ~350,000 | ~350,000 |
|  | Number of MB in incubation | ~750,000 | ~2,250,000 | ~2,250,000 |
|  | Antibody<br>functionalization/MB | 40,000/Bead | 140,000/Bead | 140,000/Bead |
| | Filtration | Salivette cotton<br>filter | N/A | 10 $\mu\text{m}$ PE filter |
|  | Reaction chamber | Centrifuge tube | Centrifuge tube | Extraction tube |
| Reader | Recording Device | Digital camera | Digital camera | Coin microscope |
|  | Device weight | ~3.5 kg | ~3.5 kg | ~1 kg |
|  | Device size (L×W×H) | 50 cm×20 cm×22<br>cm | 50 cm×20 cm×22<br>cm | 22 cm×17 cm×26 cm |
| Protocol | Total Turnaround time | 55 minutes | 6.5 minutes | 3.5 minutes |
|  | Incubation time | 30 minutes | 30 s | 15-20 s |
|  | Sampling method | Micropipette | Micropipette | Swab |
| | Specimen size | 5 $\mu\text{L}$ | 5 $\mu\text{L}$ (mock saliva) | ~94 $\mu\text{L}$ |
| | Incubation volume | 70 $\mu\text{L}$ | 70 $\mu\text{L}$ | 330 $\mu\text{L}$ |
|  | Counting method | Manual count | Manual count | MATLAB script |
| Results | BAM formation time constant | 8 min | 0.5 min | 4.4 min |
|  | Capture & detection<br>efficiency | ~85% | 43%, Fig. 3F. | ~9.2% Fig. 7C |
| | Blank BAM counts in mimic | 12.5 ± 8.8 | 4 ± 1.4 | 6.8 ± 3.2, Fig. 7C |
| | BAM counts in PCR -ve<br>saliva | 18.6 ± 10.4 | not performed | 14.9 ± 9.4, Fig. 6B |
| | LOD (N-protein molecules<br>per specimen) based on<br>capture & detection<br>efficiency and background<br>noise | 37 in 5 $\mu\text{L}$ saliva | 10-22 in 5 $\mu\text{L}$<br>saliva mimic | 1610 in 94 $\mu\text{L}$ saliva |
|  | LOD in fg/mL | 0.58 | 0.16 | 1.3 |

### 2.1 Iteration 1: Selecting larger microbubbles and reducing incubation, magnetic separation, and recording times

The original 55-minute protocol required approximately 4 minutes to extract and filter saliva using a Salivette swab, 30 minutes to mix the saliva with buoyant microbubbles and magnetic microspheres on a rotary mixer, 15 minutes for magnetic separation, and 6 minutes for signal recording. In Iteration 1, we selected larger microbubbles that rose more rapidly, allowing us to reduce the incubation time to ∼30 s, the magnetic separation time to ∼2 minutes, and the recording time to ∼30 s.

#### Selecting large microbubbles to reduce tine to separate BAM complex from free microbubbles

Magnetic separation was the second-longest step (15 minutes). A magnet rapidly pulls the BAM complexes to the bottom of the cuvette, while unbound microbubbles gradually float upwards, leaving a clear background above the magnetic particles in which to count rising BAM complexes after magnet removal. We reasoned that selecting larger, faster-rising microbubbles would reduce the required magnetic separation time.

We enriched larger, faster-rising microbubbles with five rounds of syringe-based separation, collecting microbubbles that rose quickly while discarding slower ones (see Methods Section). Supplementary Figure 1 shows photos of microbubbles separating in the syringe. This separation produced a microbubble population with diameters increasing from approximately 16 to 27 µm (Figure 3A and Supplementary Excel Sheet 1) and about 8x faster rising speed (Figure 3B). The original microbubble speed distribution has a Gaussian fraction with a mean of ∼33 µm/s, and a long tail of faster microbubbles, while the enriched fraction can be fit to a Gaussian with a mean of 248 µm/s. The increased speed arises from a combination of a larger buoyancy force from the increased volume and lower effective density (an average of 0.76 g/cm^3^ for the unenriched microbubbles, and 0.38 g/cm^3^ for the larger microbubbles, estimated from average microbubble radius, average velocity, and fluid viscosity as explained in Methods Section 4.4). Removing the smaller microbubbles also reduced the total number collected by approximately sixfold, which reduces BAM formation rate. To compensate, we concentrated the larger microbubbles ∼2.75-fold, yielding about half as many microbubbles/mL, though each microbubble has a larger surface area and rises faster.

**Figure 3:**
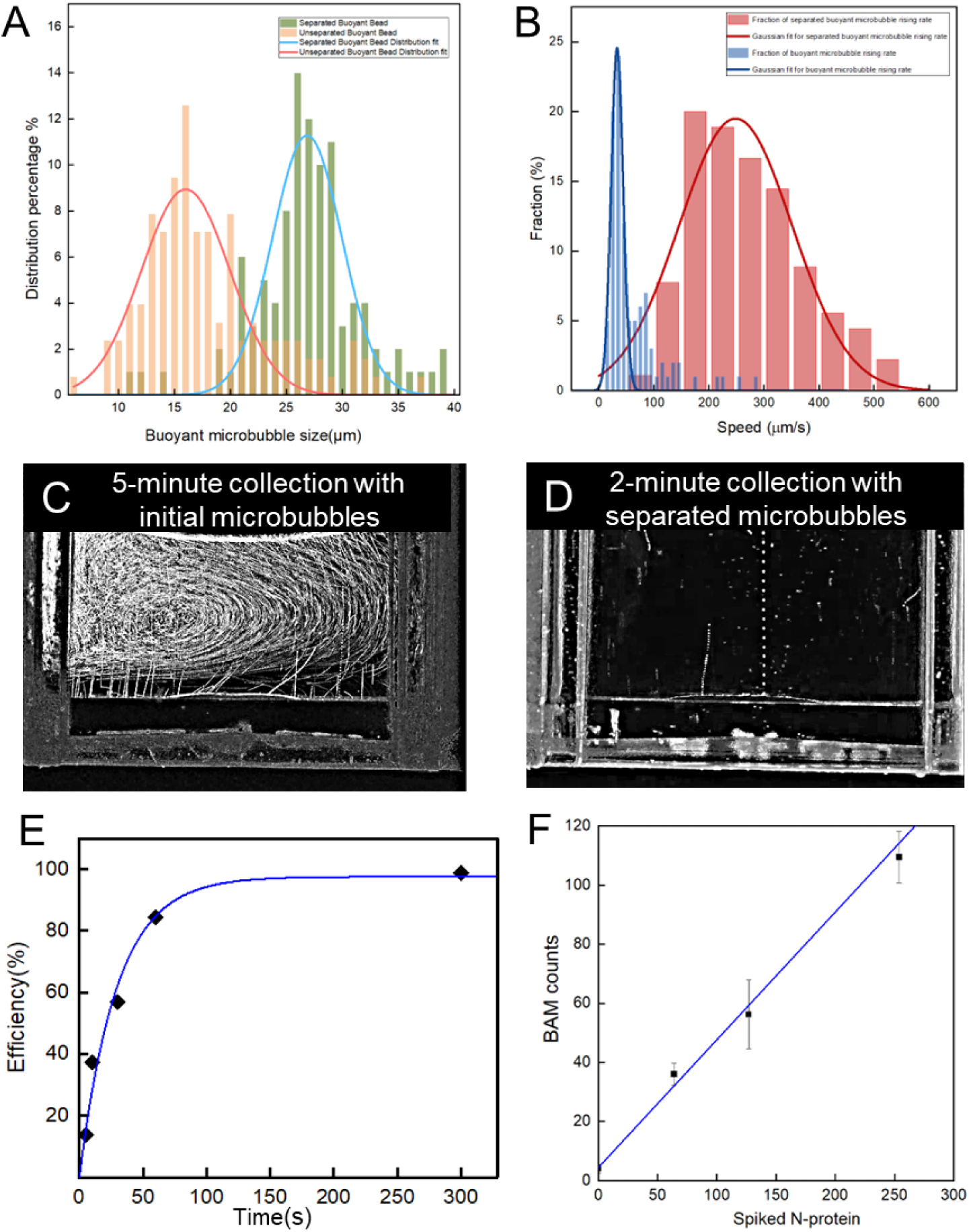
Enriched microbubble characterization. **A)** Histogram comparing speed distribution for original and large/fast enriched microbubbles. **B)** Size histogram for original and enriched microbubbles, with Gaussian regression. **C)** Non-specifically bound BAM tracks from a blank specimen using original microbubbles after a 5-minute collection. The swirls are from unbound/non-magnetic microbubbles caught in convective vortices. **D)** Non-specific BAM tracks from a blank specimen using enriched microbubbles after a 2-minute collection. **E)** Specific BAM assay capture &detection efficiency vs. incubation period, fit to an exponential with τ=32 s**. F)** Calibration curve for 0, 0.25 fg/mL, 0.5 fg/mL and 1 fg/mL N-protein spiked into 10% mock saliva, using 30 s incubation and 2 minutes magnetic separation. Datasets for Fig 3A, B, are in Supporting Excel Sheet 1, and datasets for 3E, F are in Supporting Tables 1 and 2, respectively.

A magnet pulled BAM complexes with larger microbubbles to the bottom of the cuvette within 20 seconds, while free microbubbles spontaneously floated upward and separated over minutes. The larger, faster-rising microbubbles allowed faster (2-minute) separation. Figure 3C shows BAM tracks from an analyte-free specimen after a 5-minute separation using the original microbubbles. This was below the usual 15 minutes and, consequently, showed significant background from unbound microbubbles swirling in vortices near the top of the cuvette. By contrast, Figure 3D shows a clear background after separating larger microbubbles for only 2 minutes. The larger microbubbles also generated fewer non-specific binding tracks (2 vs. 29).

#### Reducing incubation time

To increase the BAM complex formation rate, we increased the antibody concentration functionalizing the microbubbles and magnetic microspheres each by 10-fold and increased the number of magnetic microspheres in the BAM reaction mixture by 3-fold (Table 1). We counted the BAM complexes formed vs. incubation time with 10 µL of 1 fg/mL N-protein spiked into 10% simulated saliva (127 N-protein molecules/specimen on average), plus 30 µL each of buoyant and magnetic microspheres. Long incubation periods generated high capture and detection efficiency (97%, see Figure 3E) and the BAM formation time constant, *τ*, was just 32 seconds, 16x shorter than the original method.^16^ This enabled us to reduce the incubation time by 30x, from 15 minutes to 30 s, albeit with reduced capture efficiency.

To generate a calibration curve, we spiked varying N-protein concentrations into 10 µL artificial saliva and ran a BAM assay as described in the Methods Section. The BAM counts increased linearly with concentration (Fig. 3F). The slope was 0.43 counts/N-protein (43% capture & detection efficiency). For comparison, the response-time test (Fig. 3E) gave 56% efficiency at 30s, and the regression gave 63%, indicating efficient capture across experiments, with some variability in mixing and transfer times. The background from non-specifically bound complexes was 4.0 ± 1.4, which is lower with original-sized microbubbles (12.5 ± 8.8),^16^ attributed in part to opposing buoyant and magnetic forces disrupting weakly interacting complexes. The analytical limit of detection (LOD) was calculated using the formula 3s_y_/m, where s_y_ is the noise on the negative specimens and m is the calibration curve slope. Using s_y_=1.4 counts, and m = 0.43 BAM counts/N-protein, the LOD would be 10 molecules/specimen, but the 1.4 is unusually low (less than shot noise), and using the more conservative standard deviation of 3.2 counts (from pooled data in Iteration 2) gives an LOD of 22 N-proteins/specimen, improved from 37 N-proteins with the original method^16^.

#### Reducing video recording time

We reduced the video recording duration from the original protocol’s excessive ∼6 minutes to around 30 s. After magnet removal, it took 8-10 s for the fastest BAM complexes to rise to the surface, and users found 30 s was sufficient to remove the magnet and complete recording without feeling rushed.

### 2.2 Iteration 2: Improving instrumentation and protocol

Building on the results obtained with larger, faster-rising microbubbles, we further improved the assay for POC deployment across three areas: instrumentation, automated particle counting, sample handling and filtration. With the revised sample handling and filtration step, the total protocol time fell to 3.5 minutes.

#### Improving instrumentation

To transition from a benchtop assay to a rapid POC format, the bulky 3.5 kg laboratory setup (50×20×22 cm) was replaced with a 1.0 kg portable device (22×17×26 cm), see Figure 4. The new used a digital coin microscope with an integrated display, a cuvette and flashlight holder, and a magnetic separation module. A mirror redirected the image from the cuvette’s side to the upright microscope, enabling the device to operate in a compact vertical configuration. The redesigned device was configured to view a single cuvette at a time (unlike the previous 2-cuvette design). The manufacturing cost for off-the-shelf components plus a 3D-printed cuvette holder totaled $79.27 per unit (with further reductions possible through optimization and bulk production), while consumable materials cost $1.37 per test (Supplementary Table 3) and would be reduced if purchased at volume.

**Figure 4.**
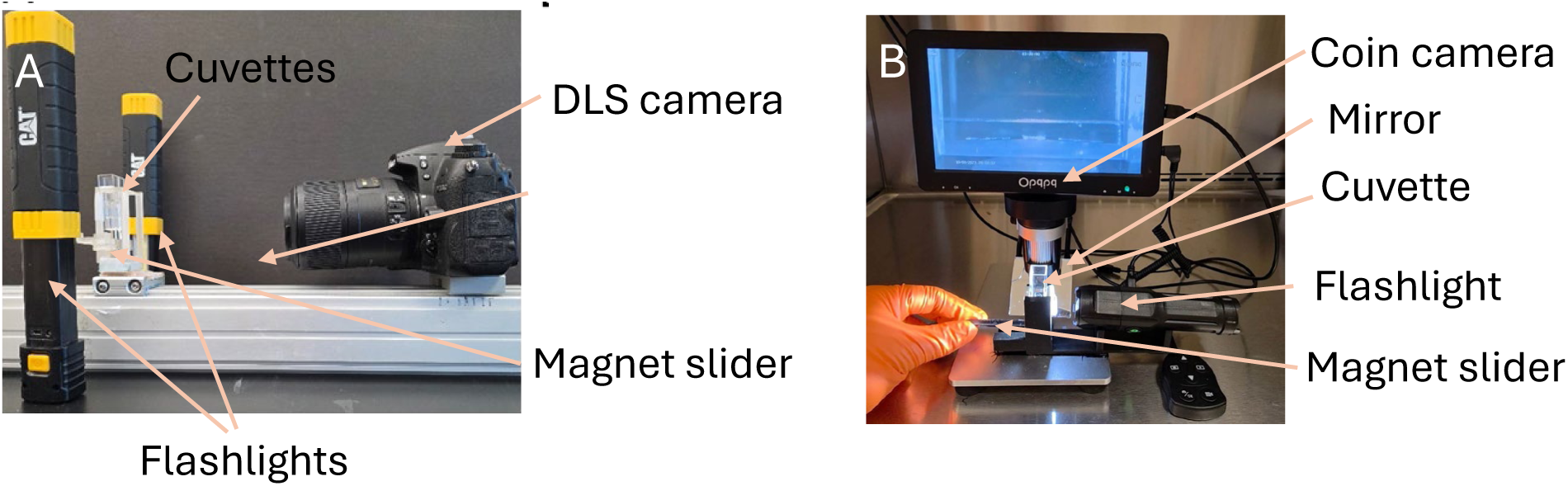
Setup photos. **A)** Original setup used in Ref ^7^ and Iteration 1. **B)** POC setup used in Iteration 2.

#### Improving BAM complex counting scripts

To simplify the BAM counting analysis, we developed a semi-automated MATLAB-based counting script that counts bright BAM complexes in a key frame 8-10 s after magnet removal (see Methods Section). Manual counting yielded a 4.9% capture efficiency, with an experimental zero-concentration background value from 9 trials of 16.7 ±9.3, while the semi-automated counting yielded a capture rate of 5.8% and an experimental zero-concentration background of 18.9 ±12.0 (Supplementary Figure 2 and Supplementary Table 4). Although the semi-automated method produced slightly higher BAM counts, the two approaches showed strong agreement at lower counts (<∼250). Additionally, semi-automation enabled quantification through 250–900 particles, when manual counting was impractical. This simplified analysis while preserving measurement consistency.

#### Simplifying sample handling and filtration

The materials were reconfigured to simplify sample handling (Table 1): The original laboratory protocol used a Salivette swab, a centrifuge to extract the saliva into a centrifuge tube, micropipettes to transfer fluid, and a 30-minute incubation on a rotary mixer. Iteration 1 reduced the incubation time but retained the other steps. In Iteration 2, the saliva was extracted from a swab and filtered in squeezable extraction tube with a 10 µm filter cap and then mixed with buoyant and magnetic microspheres in another squeezable tube. This avoided the need for a centrifuge, micropipette, and rotary mixer.

We also tried compressing a Salivette oral swab in a syringe as an alternative to using the 10 µm filter (Supplementary Figures 3 and 4). We found that the 10 μm filter was more user-friendly and showed less background variation, especially in patient saliva (Supplementary Figure 4, Supplementary Table 5). This was likely because the Salivette was designed primarily to contain and transport saliva, resulting in imperfect trapping of particulates and cells that could induce nonspecific binding. Therefore, all later studies used this 10 µm filter.

### 2.3 Results in Artificial Saliva

To analyze the effectiveness of the Iteration 2 POC assay, we first measured BAM counts vs. incubation time (5-1800 s) with 2.5 fg/mL N-protein in simulated saliva. The data fit well to an exponential regression (Figure 5A) with a 77% maximum capture & detection efficiency a 268 s (∼4.5 minute) 1/e time constant.

**Figure 5:**
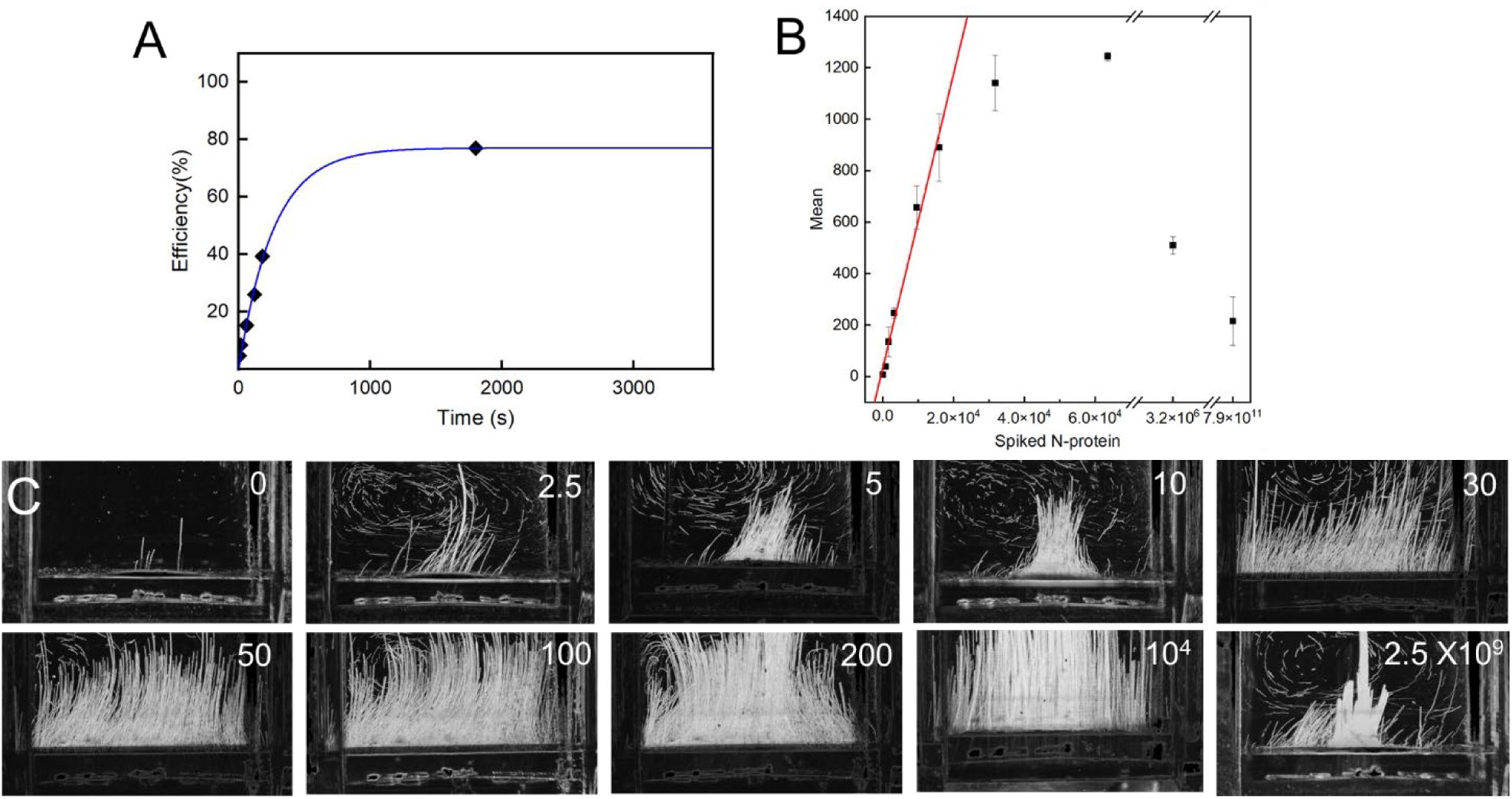
Characterizing BAM POC assay. **A)** BAM POC system capture efficiency vs. incubation period, fit to an exponential with τ=263 s. **B)** Calibration curve (0 fg/mL to 2.5 μg/mL N-protein, corresponding to 0 to 7.9×10^11^ molecules/specimen). The red line is the calibration curve fit to the low-concentration linear range of the data. Note two breaks in the x-axis for higher concentration values. **C)** Representative tracks for calibration data in B (0 fg/mL to 2.5×10^9^ fg/mL N-protein). Datasets for 5A and 5B are in Supporting Tables 6 and 7, respectively.

We also characterized the 30-squeezing cycle incubation (15-20 s) response across a broad N-protein concentration range, 0-2.5 µg/mL corresponding to 0-3×10^13^ N-protein molecules/mL. Assuming ∼500 N-proteins/virion,^16^ this corresponds to 0-6.4×10^10^ RNA cp/mL, covering the clinical range.

In the low concentration range (0 fg/mL to 200 fg/mL, i.e., 0 to 63,450 molecules added, equivalent to 0-5 RNA cp/µL), the signal increased with concentration (Figure 5B), with a slope of 0.075 counts/N-protein up to 10 fg/mL and 0.057 up to 50 fg/mL (890 counts). In the 100 fg/mL to 200 fg/mL range, the signal continued to increase slowly, eventually reaching 1244 counts. This plateau indicates that the counting algorithm saturates when the number of BAM complexes exceeds the field of view’s counting capacity.

We also ran the assay at high N-protein concentrations to ensure it remained positive. The highest tested concentration was (2.5 μg/mL), equivalent to the upper clinical range (6.4×10^7^ RNA cp/µL). At this concentration, the signal fell to 216 ±94 BAM counts, indicating a hook effect (also called prozone effect). Here, the extreme excess of antigen leads to cross-linking of large aggregates, which the counting script misinterprets as single BAM complexes. For quantification, we would need to dilute the specimen until it reaches the linear region, as we demonstrated in our earlier paper.^16^ However, this was unnecessary for a rapid yes/no determination; we observed positive results (well above the background of 7.3 counts) for the clinically-relevant concentration range from 1 fg/mL to 2.5 µg/mL (9-orders of magnitude).

### 2.4 Lab-based Results in Patient Saliva

We evaluated the Iteration 2 assay in patient saliva in a laboratory setting, beginning with a small-scale study of nine patient samples (four PCR negatives, two indeterminates with C_t_>33.5 equivalent to <1 cp/µL, and three positives) tested in triplicate, with the tester blinded to the specimen’s status. We observed clear differentiation: negative specimens yielded 6.7±2.8 BAM counts, indeterminate specimens yielded 62±35, and positive samples yielded 156±69 (Figure 6A). The BAM POC test threshold was set as 44 (halfway between the highest negative value and the lowest positive value in Fig 6A).

**Figure 6:**
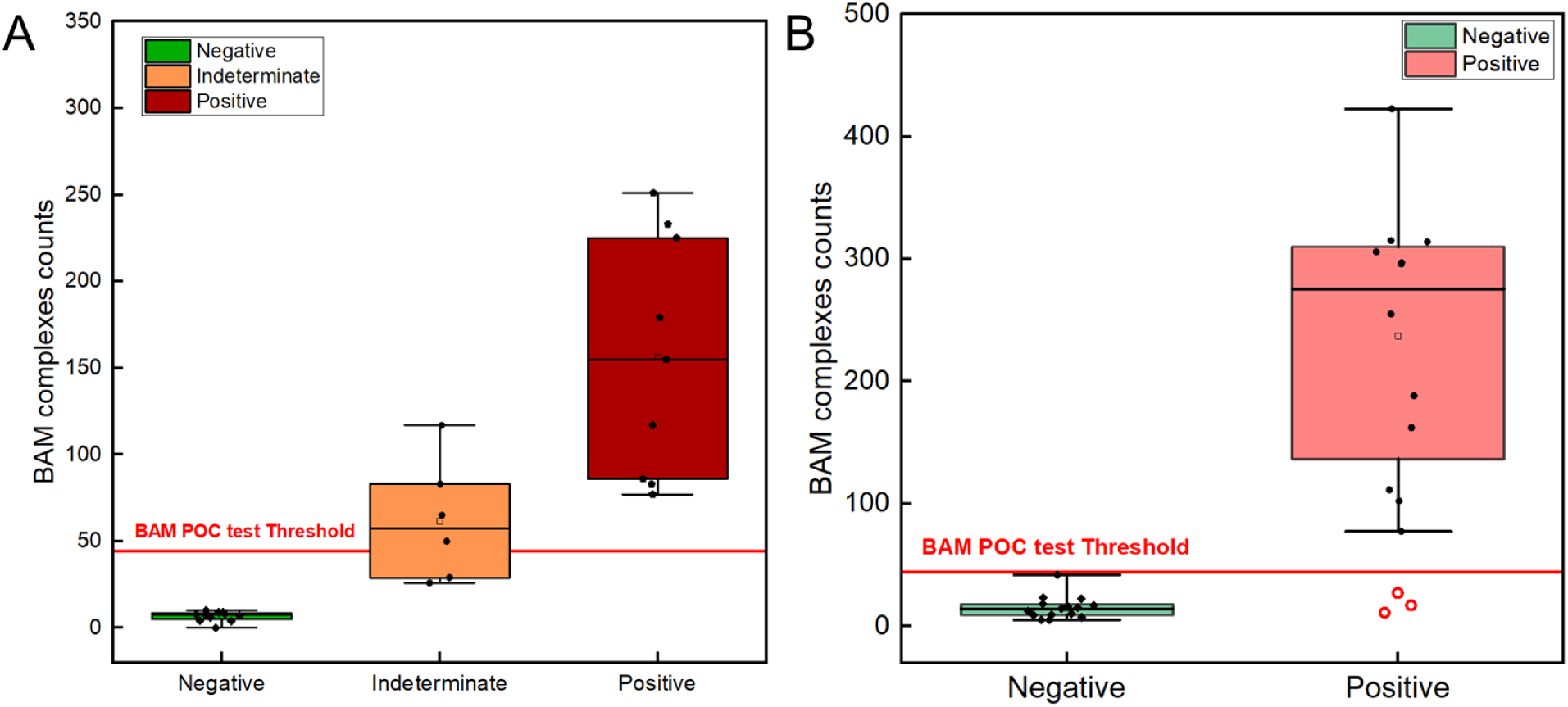
Performance validation of the BAM assay using patient saliva samples in a blinded study. **A)** Pilot blinded test results for patient saliva samples, n_positive_=3; n_negative_ =4; n_indeterminate_=2. Negative mean=6.7; standard deviation=2.8. Positive mean=156.2; standard deviation=69.1; indeterminate mean=61.7; standard deviation=34.7. BAM POC test threshold set as 44. **B)** Box plot for patient saliva sample blind test positive n=15; negative n=15; no indeterminate specimens. Negative mean=14.9; standard deviation 9.3. Positive mean=237; standard deviation=107. Datasets for A and B are in Supplementary Tables 8 and 9, respectively.

Subsequently, we performed BAM tests on a cohort of 15 PCR-positive and 15 PCR-negative patient saliva specimens, with status blinded to the user. The positive specimens had PCR C_t_ from 13.8 to 32.5, corresponding to 810,000 to 2 cp/µL (Supplementary Table 9). The negative specimens gave 14.9 ± 9.3 BAM counts, while the positive specimens gave 237 ±107 BAM counts (Figure 6B).

One negative value was unusually high (42 BAM counts versus 13.0 ±5.8 for the 14 other negatives), and just below the threshold of 44 BAM counts. This sample had a slightly yellow tint, suggesting an unknown interferant or possibly a weakly positive case reading negative on PCR (repeated PCR on a remaining 24 µL of specimen was still negative, but this was insufficient to rule out a weak positive <∼0.05 virion/µL).

Three false negatives were also observed from specimens with PCR C_t_ values of 19.6, 21.7, and 30.5 (corresponding to ∼14,500, 3,400, and 8 cp/µL, respectively). We retested these specimens and diluted them serially by 10x factors to reduce matrix effect (e.g., from interfering antibodies or other substances in the saliva). All three specimens reproduced a false negative at the original saliva concentration (Supplementary Table 10). However, with sufficient dilution, two of the specimens gave positive results: At 10,000x dilution, the C_t_=19.6 specimen became clearly positive (115 counts); and the C_t_ 21.7 became clearly positive (143 counts) at 100x dilution. The third specimen did not become positive at up to 10,000x (the highest dilution tested, where the value started to fall), but it was relatively diluted to begin with (C_t_=30.5, corresponding to about 8 RNA cp/µL). Notwithstanding these discrepancies, the positive results confirmed that we could detect low viral concentrations (as low as 2 cp/µL) with our rapid protocol, with no false negatives (although one value was near threshold).

### 2.5 Point-of-care studies

We simulated POC use of the Iteration 2 BAM assay on a hospital ED workbench located 40 miles from Clemson University (Figure 7A) and on the trunk of an emergency response vehicle (Figure 6B). These tests were performed by both an ED technician and a community paramedic. Standards in simulated saliva (0-10 fg/mL N-protein) confirmed that all operators properly operated the assay. The capture &detection efficiencies across the trails were: 7.5% for lab measurements by a graduate student (this was the same data as in Fig 5B), 10.8% for the lab measurements by the ED technician, 10.1% for the ED lab measurements by the ED technician, and 8.4% for the paramedic in the trunk of an emergency response vehicle (Figure 6C). Across the pooled data, the slope was m=0.092 BAM counts/molecule (9.2% capture & detection efficiency in the Triton-X 100 diluted saliva, or 1.75% of original swab), and the zero-concentration standard deviation was 3.2 BAM counts (Supplementary Figure 5, Supplementary Table 11), yielding an analytical LOD (3s_y_/m) of 106 molecules, equivalent to 0.3 fg/mL in 94 µL original saliva sample before lysis. These results showed all operators could run the test quickly and easily with excellent LODs and semi-quantitative results in artificial saliva.

**Figure 7:**
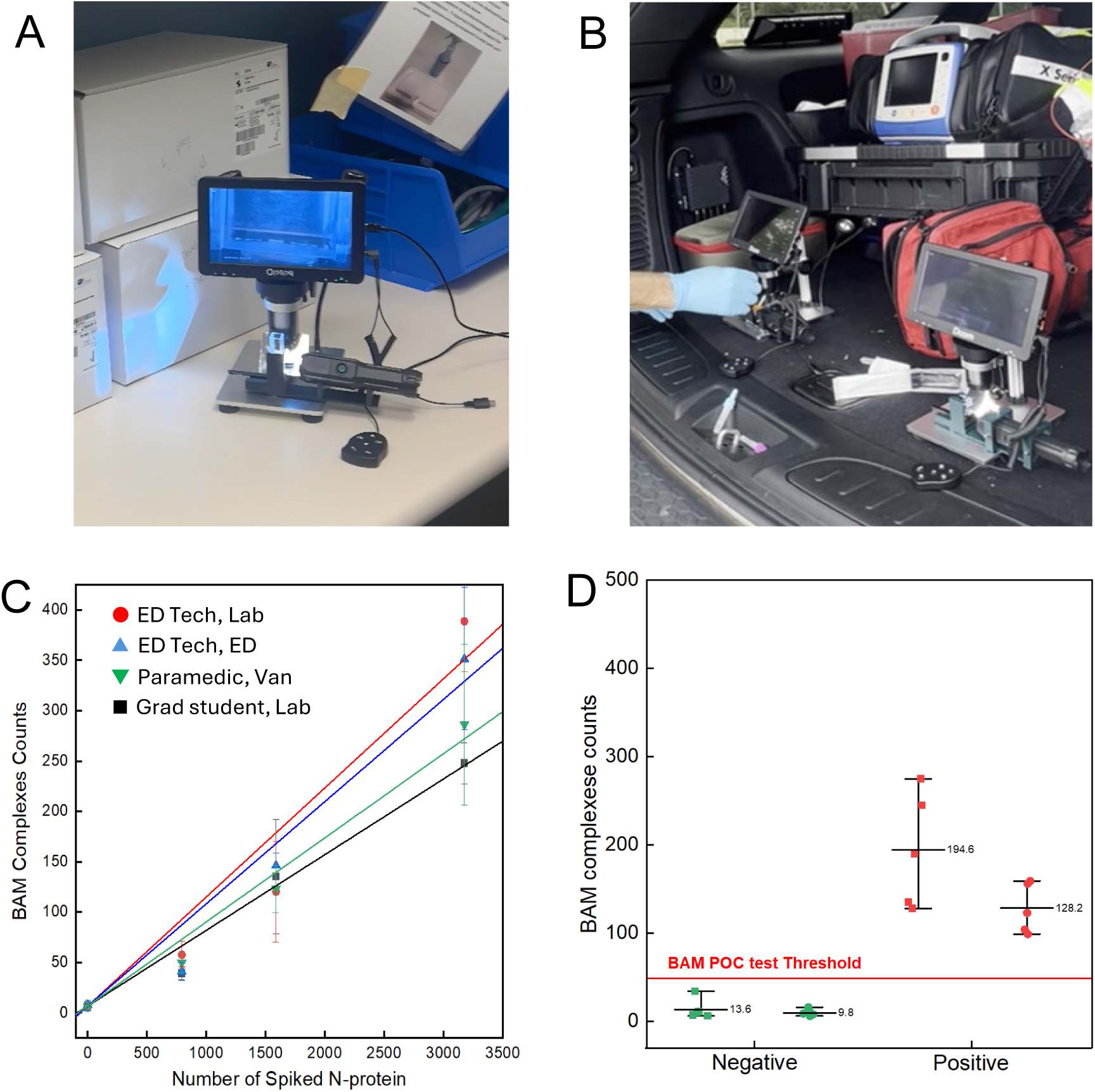
Clinical validation and inter-operator agreement for the BAM POC assay. **A)** Photo of BAM POC setup on a hospital ED bench. **B)** Photo of two BAM POC setups in a parked van trunk. **C)** Calibration standards in simulated saliva (0, 2.5, 5, and 10 fg/mL N-protein) measured by three different operators across varied experimental environments (graduate student in research lab, ED technician in research lab, ED technician in hospital ED assay room, and community paramedic in parked van). The pooled zero-concentration values were 6.8±3.2 BAM counts. **D)** Inter-operator comparison of BAM counts for n=5 negative and n=5 positive specimens measured by two operators, ED technician and paramedic on the back of a parked van. The negative sample means were 13.4 (SD=11.6) and 9.8 (SD=3.8); the positive sample means were 194.6 (SD=64.2) and 128.2 (SD=28.2). The BAM POC test threshold was set at 44 BAM counts. Datasets for C and D in Supplementary Tables 11 and 9, respectively.

We next assessed whether the operators could properly test patient saliva. Five positive and five negative saliva specimens were randomly selected from the cohort previously lab-tested in Figure 6B. To minimize biosafety concerns, prior to transport, the saliva was mixed with Triton-X 100 to lyse the virus and a protease inhibitor cocktail and stored on ice, conditions that preserved the N-protein for at least 8 hours (Supplementary Figure 6). The ED technician and paramedic tested each specimen once on the back of a parked van, blinded to specimen status (Figure 7D). The negative specimens for the two operators averaged 13.4 and 9.8 BAM counts, while the positive specimens averaged 194.6 and 128.2, with all n=10 PCR-positive trials above threshold, and all n=10 PCR-negative trials below threshold. Overall, the study showed that the system operated well in the field, confirming its resilience to environmental factors and its operational simplicity.

## 3. Discussion

The BAM immunoassay demonstrates a unique combination of sensitivity, simplicity, affordability and speed: 1.3 fg/mL LOD in patient saliva, run in a parked vehicle within 3.5 minutes (Table 1). This LOD corresponds to about 1610 N-proteins or 3.4 virions per 94 µL swab, assuming about 470 N-proteins/virion (based on our previous BAM study^16^ and consistent with electron microscopy^17^). This is comparable to q-RT-PCR (which can detect down to 1 RNA molecule but often use small volumes). The BAM POC assay meets the WHO’s 2020 TPP for minimum for both optimal POC LOD (10^3^ cp/uL) and minimum diagnostic LOD (1 cp/µL, corresponding to approximately 38 fg/mL).^5^ Indeed, the assay could detect N-protein in samples classified as indeterminate by q-RT-PCR at the Clemson Research and Education in Disease Diagnosis and Intervention Laboratory (REDDI) Lab (<∼1 cp/µL)^18^ (Figure 6A).

Table 2 compares our BAM assay with two commercial ultra-sensitive SARS-CoV-2 N-protein immunoassays: Quanterix Simoa (a magnetically separated single-molecule counting enzyme-linked sandwich immunoassay) and Mesoscale Discovery S-plex (an electrochemiluminescence sandwich assay). Only the custom-developed Simoa and BAM assays have limits of detection sufficient to see <1 cp/µL. Uniquely, the BAM assay combines ultrasensitivity with portability, simplicity, and rapid time-to-result, and low cost (Supplementary Table 3).

**Table 2.** Comparison of BAM POC assays with two commercial ultrasensitive (though not portable) SARS-CoV-2 N-protein assays.

| Parameter / Feature | Quanterix Simoa | S-plex | BAM POC Assay |
| --- | --- | --- | --- |
| Reference | Olsen et al.<sup>19</sup> | Ren et al.<sup>20</sup> | This publication |
| LOD | 99 fg/mL commercial; 20 fg/mL in-house | 160 fg/mL (2.5 s<sub>y</sub>/m) | 1.3 fg/mL |
| Turnaround time | 2.5 h + transport | 8 h + transport | 3.5 minutes |
| Dimensions (cm) (and volume, L) | 135 × 60 × 160 (1296 L) | 68×77×86 (450 L) | 22 × 17 × 26 (9.7 L) |
| Supply | power supply | power supply | power/battery supply |
| Throughput (trials/run) | 96 | 2000 | 1 |

Although BAM assays are not as simple, small, and equipment-free as LFAs, they are simple enough for POC use and can be further automated in future. Most commercial LFAs have LODs in the pg/mL-ng/mL range, which result in up to ∼50% false negatives in asymptomatic patients and ∼30% in symptomatic patients.^4,13^ While LFAs can detect highly infectious patients, patients with viral loads below their LOD can still be infectious (or may become so soon), especially with prolonged close contact, such as in families.^2,7,8^ Ultrasensitive diagnostics such as BAM could therefore enable earlier detection, supporting faster clinical intervention and enhanced transmission control. Additionally, the saliva used in this BAM assay is more convenient than nasal swabs used in most LFAs and typically becomes PCR-positive earlier (although with significant patient-to-patient variation).^3,21^ Finally, the BAM assay provides results in 3.5 minutes, while most LFAs take 15-30 minutes to fully develop, which is useful in emergency medicine and mobile health. The speed and sensitivity could also open new applications to explore in the future, for example, to rapidly detect airborne viruses.

The remarkable analytical LOD can be attributed to bright single-particle detection, rapid and efficient analyte capture, and low background. The analytical LOD is defined as the lowest analyte concentration that can be reliably distinguished from the blank. It is commonly given by Equation 1:

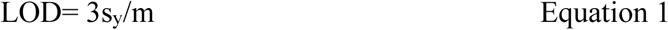

where s_y_ is the standard deviation of the signal from negative specimens and m is the low-concentration calibration curve slope (signal per analyte molecule or concentration). In an immunoassay, the signal per analyte molecule, m, is given by the fraction of analyte molecules labeled (η) multiplied by the signal per label (I_label_). In conventional assays, including LFAs, the LOD can be improved by increasing I_label_. LFAs typically employ 40 nm gold nanosphere labels with ∼10^4^-fold higher absorption cross sections than typical visible dye molecules to increase I_label_.^22^ Even so, around 10^7^ nanosphere labels must bind to an assay strip to become visible to the naked eye.^23^ By contrast, scattering from a single 20 µm microbubble is observable to the naked eye because it has a 10^5^-fold larger surface area and a 10^8^-fold larger volume than a 40 nm gold nanosphere; the scattering also provides a bright signal against a dark background, and the rising motion provides additional specificity. However, 20 µm labels cannot be used in LFAs because they would fit through the nitrocellulose membrane pores, would settle or rapidly,^24^ would experience high torques from shear flow,^25,26^ their slow diffusion rate would reduce capture efficiency,^24^ and their higher surface area would make them prone to non-specific binding from rare defects in surface coverage/blocking.^24^ The BAM format avoids these large-label issues by speeding the reaction using high concentrations of microbubbles and buoyant mixing, and reducing non-specific binding via opposing magnetic and buoyant forces.

When labels can be individually counted, the LOD becomes independent of I_lalbel_ because brighter and dimmer labels count equally. s_y_ then becomes the variation in average background counts, δB (including shot noise and specimen variation), and m = η (capture-and-detection efficiency, 0-100% counts/molecule) (Equation 2).

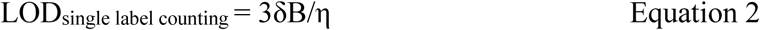

For Iteration 1, η was ∼97% after five minutes, or 43% with 15 s incubation (Figure 3). Since the simulated saliva background was 4 counts, this was approaching the theoretical limit of counting every molecule (100% capture-and-detection with 0 background). For context, LFAs often have label capture efficiencies of <1%,^23^ although this can be increased by slowing fluid flow through the test zone.^27^ Meanwhile, Simoa assays usually employ long incubation times (e.g., 35 min) to allow reactions to complete. Although long incubation times generate high analyte capture efficiencies, only ∼5% of magnetic microspheres are loaded into microwells for detection, limiting η overall.^28^ They also typically have higher background counts (and variation therein) than BAM assays, raising (worsening) the LOD. Also, the setups are large, expensive, and operated in centralized labs, with significant shipping and setup times and run times of 80 minutes for a single test or 2.5 hours for a 96-well plate.^11^

Since our primary goal was POC detection, in Iteration 2, we sacrificed some marginal capture efficiency/sensitivity for significant speed and user-friendliness improvements. We used a 94 uL swab to collect saliva, lysed it in Triton-X solution, which diluted the specimen, mixed it with buoyant and magnetic microspheres, using a large enough incubation volume to pour into the cuvette and read without adding more buffer. The diluted incubation solution reduced the reaction rate compared to Iteration 1 by ∼9-fold, increasing the time constant from 29 s to 263 s (Figures 3E and 5A, respectively). This decreased the capture efficiency for the 15-20 s incubation to ∼9.2% in pooled diluted saliva (Figure 7C). Together with the Triton-X dilution, these changes reduced the overall η to ∼1.75% of the molecules in the original 94 µL swab. Artificial saliva typically shows backgrounds largely limited by shot noise, whereas patient saliva shows both a higher background and greater variability than can be accounted for by shot noise, implying some specimen-dependent interference. Nonetheless, the POC assay sampled a larger initial volume (94 vs. 5 µL) containing more molecules and reduced the non-specific binding background by using larger microbubbles and 10 µm filtration. Overall, the POC assay LOD was 1.3 fg/mL, 2.5-fold worse than our original 55-minute protocol, yet still well below the WHO diagnostic TPP.

While the results show an extraordinary combination of speed and ultrasensitivity in a POC assay, the protocol, instrumentation, and reagents remain far from optimized. Several limitations and opportunities remain: First, above ∼900 BAM complexes, the counting became saturated, preventing quantification. Nonetheless, the assay demonstrated qualitative (yes/no) detection over nine orders of magnitude (1 fg/mL–2.5 µg/mL), which is sufficient for clinical decision-making. If required, quantitative performance can be extended through specimen dilution, as we previously demonstrated,^16^ and by refining analysis scripts to better account for particle overlap and aggregation.

Second, Figure 6B shows three false-negative results. These were likely from competitive interference from endogenous antibodies in the saliva or other matrix effects, since dilution increased the signal. To overcome such matrix effects in future, we could pre-treat the saliva to remove existing antibodies and other potential interferants, preferably prior to viral lysis, and/or further dilute clinical samples, albeit with some sensitivity loss. Conversely, one negative specimen had a much higher BAM count than the others (42 in Fig 6B, and 34 and 10 by the two healthcare workers in Figure 7D). An unknown interferent may have elevated the non-specific binding, or the BAM assay may have detected low levels of N-protein not observed by PCR. More studies will be needed on a larger pool of specimens to understand the causes and frequencies of false-positive and high-negative results.

Third, in terms of protocol, reagents, and instrumentation, manual operation introduces potential operator-dependent variability in quantification. Automating processes such as mixing could reduce this variation, though it would make the instrumentation or disposables more complex. Additionally, the instrument runs only one test at a time. In principle, BAM systems could be modified to measure multiple specimens by viewing several channels simultaneously or by sequentially interrogating specimens. However, the current instrument prioritizes high-speed, ultra-sensitivity, and POC deployment over the batch-processing capacity of laboratory-based systems. We must also work on long-term functionalization and storage of microbubbles and magnetic microspheres. This could be achieved by covalent particle functionalization (in place of biotin-streptavidin linkages) and subsequent lyophilization in a sucrose or similar matrix, as is done for LFAs. Additionally, future iterations should incorporate internal procedural validations. For example, colored solutions could help ensure reagents were added and at the proper concentration. The system could validate reagent integrity and protocol performance by splitting the cuvette and running a parallel control sample alongside the patient sample. Concurrently, data management capabilities should be enhanced to perform on-site data processing and generate automated diagnostic reports.

Fourth, this study emphasized practical implementation. Additional studies will be needed to elucidate fundamental questions about reaction rates for specific and non-specific binding for each step of BAM complex formation, as well as dissociation rates under magnetic and buoyancy forces. This will be important for extending the assay to other analytes with different antibody-antigen reaction kinetics under force, as well as analytes with multiple binding sites such as cells. Such modeling requires significant effort as the system is complex, including magnetic chaining in magnetic fields, fluidic and magnetic interactions between magnetic and buoyant microspheres, and a combination of antibody-antigen interactions plus surface interactions between microspheres.

Finally, regulatory agencies such as the FDA would need to approve BAM assays before clinical use. The device, protocols, and quality systems would need to be finalized and tested in a large-scale, multi-site clinical study to demonstrate substantial equivalence to current gold-standard assays. A CLIA Waiver (Clinical Laboratory Improvement Amendments) would be needed to permit device use in non-laboratory settings such as pharmacies and mobile clinics.

In summary, although much work remains, the BAM assay represents a new class of POC diagnostics, combining rapid time-to-result with ultra-sensitivity. Its 3.5-minute turnaround time constitutes a dramatic departure from existing high-sensitivity platforms such as Simoa and S-plex. These improvements are achieved at the POC, thereby avoiding the transportation delays, cost, and logistical constraints that dominate conventional diagnostic workflows. Finally, BAM assays use low-cost devices and consumables (Supplementary Table 3), providing a simple and scalable platform for POC diagnostics.

## 4. Methods

### 4.1 Materials

The Streptavidin functionalized buoyant microbubbles (SKU:11110-000) and separation buffer (bundled package with buoyant bead) were obtained from Akadeum Life Science (Ann Arbor, MI, USA). The LodeStars streptavidin 2.7 μm magnetic microspheres (Cat: PL6727-1001) were purchased from Agilent Technologies (Santa Clara, CA, USA). A PBS solution (Gibco PBS pH 7.2, Cat: 20012027) was sourced from ThermoFisher Scientific (Rockford, IL, USA), along with the Pierce Antibody Biotinylation Kit for IP (Cat: 90407). Biotin-mPEG 5K (Cat: PLS-2054) was obtained from Creative PEGworks (Durham, NC, USA). The SARS-CoV-2 nucleocapsid antibody (Rabbit Mab, Cat: 40143-R004, 40143-R040, hereinafter referred to as Ab-004 and Ab-040) was purchased from Sino Biological (Paoli, PA, USA), while the SARS-CoV-2 nucleocapsid protein (N-Protein, a 47.3 kDa protein synthesized with a his-tag, Cat: NUN-C5227-100 µg) was acquired from Acro Biosystems (Newark, DE, USA). Triton X-100 was sourced from Alfa Aesar (Ward Hill, MA, USA), and Milli-Q water (18.2 MΩ/cm) was obtained from an ELGA Purelabflex2 system (ELGA LabWater, Woodridge, IL, USA). For sample collection, a cotton saliva swab (Salivette, REF: 51.1534) was acquired from Sarstedt AG (Nümbrecht, Germany). The extraction tube for the iHealth COVID-19 Antigen Rapid Test was supplied by iHealth Labs, Inc. (San Jose, CA, USA). The extraction tube cap with built-in 10 µm filter (Model: YGC05) was obtained from Foshan Yuyang Medical Instrument Co., Ltd (Foshan, Guangdong, China). The semi-micro cuvette (Cat: BR759115) was purchased from MilliporeSigma (Burlington, MA, USA). Halt™ Protease Inhibitor Cocktail (Lot: 3212213) and low protein-binding centrifuge tubes (Cat: 90401) were obtained from ThermoFisher Scientific (Rockford, IL, USA). The 7″ digital video microscope, from Dcorn was purchased from Amazon (Seattle, WA, US), and magnets including 2 disc magnets (Cat: D1051A, diameter 8 mm, thickness 4 mm, Neodymium strength 45) and a cylinder magnet (Cat: Cyl0164, diameter 6 mm, height 8 mm, Neodymium strength 50) were sourced from SuperMagnetMan (Pelham, AL, USA).

### 4.2 Antibody biotinylation

To biotinylate the N-protein antibodies (Ab-R004 and Ab-R040), the “Pierce Antibody Biotinylation Kit for IP” (Catalog #90407, ThermoFisher Scientific, Waltham, MA, USA) was used. This kit reacts with the amine groups on the antibodies via an NHS-PEG4-biotin linker. This step is detailed in previously published articles.^7^

### 4.3 Large buoyant microbubble enrichment

We first took a 10 mL syringe and rinsed the inner walls and the plunger with 0.1% Triton-X 100 in 1x PBS solution (Supplementary Figure 1). This rinsing minimized the adsorption of buoyant beads to the syringe and plunger during separation. 2 mL of buoyant microbubbles were then transferred into the syringe. 8 mL 0.1% Triton-X 100 in 1x PBS solution was aspirated into the syringe. The syringe was positioned with the tip facing upward and allowed to stand for 30 minutes at 4 °C to collect all buoyant beads at the plain tip end.

Next, the syringe was inverted with the tip facing downward and allowed to stand for 5 minutes at 4 °C to gather the larger buoyant beads at the plunger end. The smaller buoyant beads remained distributed within the syringe. Subsequently, the solution was expelled from the syringe into a waste container, while the larger buoyant microbubbles were retained. These beads were significantly whiter and showed a clear demarcation from the solution containing smaller buoyant beads. Finally, 0.1% Triton-X 1x PBS solution was aspirated up to the 10 mL mark of the syringe. The above steps were repeated five times until a clear solution was expelled.

The solution containing the larger buoyant beads was expelled from the syringe into a 5 mL centrifuge tube. 1 mL of 1x PBS solution was aspirated into the syringe, mixed thoroughly, and expelled into the 5 mL centrifuge tube above. This process was repeated three times to wash the buoyant beads in the syringe. The tube was then centrifuged at 400×g for 3 minutes, and the supernatant was removed. 700 µL of separation buffer provided by Akadeum was added as the storage solution. The final product was stored at 4 °C.

The microbubble density, ρ, was estimated from their average radius, r, average velocity, v, fluid viscosity, η (10^-3^ Pa-s), and gravitational constant on earth (9.8 N/kg) by balancing the buoyancy force (^4^/_3_ πr^3^ ρ g) and Stokes viscous drag (6π η r v), as ρ=^9^/_2_ η v r^--2^ g^-1^.

### 4.4 Microbubble Surface Modification

#### Buoyant Microbubble Surface Modification

20 µL of 0.05 µg/µL Ab-004 was added to a 2 mL low protein-binding centrifuge tube. 100 µL of the larger separated buoyant microbubbles was added and mixed by pipetting in and out 50 times. The mixture was incubated at room temperature for 10 minutes. Next, 100 µL of 0.01 mM mPEG 5k-biotin in 1x PBS solution was added and the mixture was pipetted in and out 50 times again. Then, 400 µL of Akadeum Life Sciences separation buffer (containing Ca2+ and Mg2+ free PBS, 2 mM EDTA, 0.5% biotin free BSA, 0.09% sodium azide, and additionally spiked with 0.1% Triton-X 100 surfactant) was added and mixed thoroughly. The sample was centrifuged at 400×g for 3 minutes. The subnatant was removed, and the washing steps were repeated two more times. The buoyant beads were stored in 400 µL 0.1% Triton-X 100 with separation buffer at 4 °C. These functionalized microbubbles were used within 24 hours of functionalization to ensure reproducible capture efficiency and low nonspecific binding.

#### Magnetic Microsphere Surface Modification

20 µL of 0.05 µg/µL Ab-040 were mixed with 36 µL of Agilent Lodestar streptavidin-functionalized magnetic beads and pipetted in and out 50 times. The mixture was incubated in a 2 mL low protein-binding centrifuge tube at room temperature for 10 minutes. Then, 100 µL of 0.01 mM mPEG 5k-biotin 1x PBS solution was added and pipetted in and out 50 times. Following this, 400 µL of separation buffer containing 0.1% Triton-X 100 was added and mixed thoroughly. The mixture was centrifuged at 400×g for 3 minutes. The supernatant was discarded and the washing steps were repeated twice more. The mixture was stored in 400 µL 0.1% Triton-X 100 with separation buffer at 4 °C. These functionalized microspheres were used within 24 hours of functionalization to ensure reproducible capture efficien8cy and low nonspecific binding.

### 4.5 Saliva

Experiments were performed in both simulated saliva and human saliva specimens.

#### Simulated saliva studies

Artificial saliva was made using the following: 15.6 mM NaCl, 16.5 mM KCl, 1.01 mM CaCl_2_, 0.361 mM MgCl_2_, 2.07 mM K_2_HPO_4_, 16.3 mM urea, and 5.0 g/L carboxymethylcellulose sodium salt in one aqueous liter.^29^

#### Human saliva collection

The saliva collection protocols were approved by the Institutional Review Boards (IRB) of Clemson University (IRB2021-0703) and Prisma Health (Pro00100731). Experiments with the saliva were performed under approved Clemson University Institutional Biosafety Committee (IBC) protocol IBC2022-0138. All study participants provided their written informed consent to provide a saliva sample. Participants provided 1-2 mL of saliva collected in a 50 mL conical tube and demographic data (age, gender, race/ethnicity) to the research study.^30^ Samples were deidentified and stripped of personal identifying information before being passed to the research team. Part of the samples (200 µL) was used for a RT-PCR clinical diagnostic test^18^ and eventual sequencing for SARS-CoV2 variant identification.^31^ For this protocol and instrument, RNA copies/µL are related to cycles to threshold (C_t_) as: RNA copies/µL = 2^(Ct–33.5)*0.947^, although the C_t_ values exhibit a trial-to-trial variation of ∼0.8 C_t_ units with larger discrepancies for dilute (high C_t_) specimens. Note: the conversion between C_t_ and concentration is protocol and threshold-dependent; thus, C_t_ values should not be directly compared across studies that use different procedures. Digital PCR would be needed for more accurate quantification.

### 4.6 Spiked saliva preparation

990 µL of PCR-negative saliva was transferred to a 2 mL low protein-binding centrifuge tube and 10 µL of 250 μg/mL N-protein solution was added. Subsequently, this spiked saliva solution was diluted with additional PCR-negative saliva to achieve the desired N-protein concentration. This solution was prepared on the day it was used and was briefly stored (<8 hours) in an ice bath.

### 4.7 Experiment Setup

As shown in Figures 4B, 7A, and 7B, the experimental setup for the BAM POC settings consists of a 7-inch digital video microscope, flashlight, a 5 cm x 5 cm mirror, magnets, and 3D-printed components: flashlight holder, cuvette holder, magnet slide, mirror holder, and mirror slide rail. The flashlight holder and mirror slide rail are integrated with the cuvette holder. The mirror can be adjusted by moving the holder back and forth on the slide rail to adjust the reflection view in the microscope. The video recorded by the coin microscope is stored on a 32 GB SD card.

### 4.8 BAM point-of-care assay protocol

Figure 8 shows the BAM POC test protocol from Iteration 2, and a more detailed Instruction Booklet is in Supplementary Figure 8, and Supplementary Figure 9 shows photographs of some of the steps.

**Figure 8:**
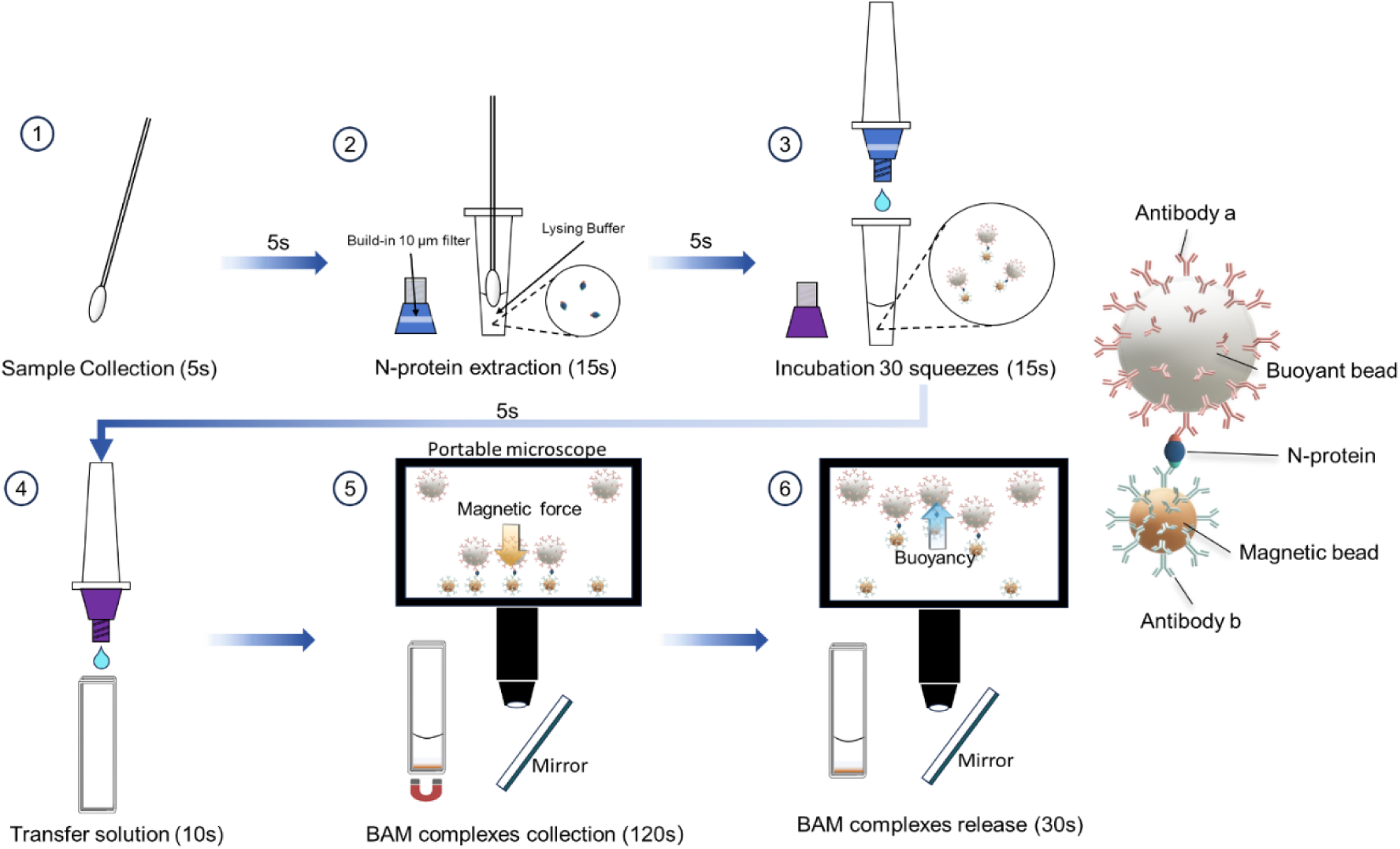
BAM POC test protocol schematic. Total assay time is 210s. Supplementary Figure 8 shows a more detailed instruction booklet.

To enhance usability, we employed two reagent tubes with different colored droppers to visually differentiate their contents. The Lysis Buffer Tube had a blue dropper equipped with a built-in 10 μm PE filter. This tube contained the lysis solution: 400 μL of 0.5% Triton-X in 1×PBS, designed for viral lysis and simultaneous sample filtration. Conversely, the probe reaction tube, identified by a purple dropper cap, is an extraction tube without an integrated filter, designated specifically for the incubation step. The probe reaction tube was pre-loaded in the laboratory with the essential detection components: 30 μL buoyant microbubbles, 30 μL magnetic microbeads, and 140 μL of 0.1% Triton-X separation buffer (0.5% BSA, 2 mM EDTA, 1 × PBS solution), serving as the designated container for BAM complex formation.

The swab was immersed in the saliva and stirred 10 times to collect the saliva solution (∼94 μL). This solution was then transferred into the lysis buffer tube, which was equipped with a blue dropper that had a built-in 10 μL PE filter. The swab was stirred in the lysis buffer tube 15 times, and as the swab was removed, the tube was squeezed to ensure as much liquid as possible remained inside. The probe reaction tube preloaded with buoyant microbubbles and magnetic microspheres was shaken briskly to disperse the particles and the cap was then removed. Three drops (∼ 131 μL) of the prepared testing saliva solution were then transferred into the probe reaction tube. The mixture was incubated and agitated by squeezing the tube 30 times (∼15 seconds) to ensure thorough mixing and the formation of BAM complexes. Subsequently, the reaction solution was transferred to a cuvette. The cuvette was placed on the cuvette holder above the magnet, which collected BAM complexes while free microbubbles floated upwards. After 2 minutes of magnetic collection, the video recording was started, and the magnet was immediately removed by sliding it in its track away from the cuvette. Magnet removal released BAM complexes, allowing them to float upward and be seen via flashlight illumination.

### 4.9 Counting BAM complexes

We used two methods to count BAM complexes in a video: Iteration 1 used a mostly manual approach and Iteration 2 used a semiautomatic approach.

The manual approach was described in our earlier publication.^16^ Briefly, ∼30 s of video (recorded at 33 frames/second) was analyzed after the magnet was removed. The maximum and minimum intensities were calculated for each pixel in the movie. Then the maximum minus minimum image was calculated. This image subtracted out any fixed background (where maximum and minimum intensities were similar throughout the movie), and highlighted regions where a bright BAM complex or microbubble passed through (where the minimum came from a time without the BAM complex in view, and the maximum occurred with it). When necessary, this image was superimposed with the first image to show the particle tracks against the background cuvette image. A series of 10 s intervals was examined, and in each interval, the number of new tracks rising from the bottom of the cuvette was counted manually.

The semi-automatic counting script was developed to simplify and expedite the BAM counting process (printed in the Supporting Information). This MATLAB script counted the bright objects of sufficient size in a selected frame of a movie. To select the appropriate frame, we manually reviewed the video after magnet removal and identified the last frame before the fastest BAM complex leaves the field of view. To avoid counting bright but stationary objects, we first calculated a background image and subtracted it. For each pixel location, the background signal was calculated as the minimum intensity in time from the movie sequence. After background subtraction, the script circles each remaining bright particle of appropriate size and intensity, and users can remove occasional extra counts from free buoyant microbubbles that are not rising from the bottom (evident in the video and lightly superimposed BAM tracks), or add rare missed BAM complexes. The larger microbubble size created bright BAM complexes with low background from unbound microbubbles, making this approach straightforward and effective, with few manual count revisions needed. The script is given in Supplementary Information.

### 4.10 Signal Stability and Storage Conditions

To evaluate the stability of the target antigen in clinical matrices during storage and transport, a time-course degradation study was performed (Supplementary Figure 7). Recombinant N-protein was spiked into PCR-negative saliva samples to a final concentration of 10 fg/mL. A baseline signal was established by testing the spiked samples immediately (t=0). The lysis buffer tubes contained lysed saliva samples and were subjected to two experimental conditions. In the first set, samples were stored without additives at 4 °C and 20 °C (room temperature). In the second set, to mitigate enzymatic degradation, 10 μL of protease inhibitor cocktail was added to each lysis buffer tube, and storage temperatures were set to 4 °C and 0 °C (ice bath). Signal retention was quantified in triplicate at 2-, 4-, and 8-hour intervals.

### 4.11 Reagent Transportation

All pre-aliquoted bottles (lysis buffer tubes and probe reaction tubes) and solutions required for field deployment testing were stored in an ice bath, transported using a Styrofoam cooler, with a transit time of 20-40 minutes.

### 4.12 Test Sites

The experiments in the research lab were conducted in a biosafety hood (Class II type A2, from Labconco, Kansas, MO, US) at the Biosystems Research Complex at Clemson University. The experiments in an ED department were conducted on saliva simulants on a desk in the Emergency Department’s Respiratory Care ABG Laboratory in Greenville Memorial Hospital.

We performed the experiments on the back of a van in the parking lot over two days, one at Oconee Memorial Hospital and a second at Prisma Health Baptist Prisma Hospital. The first day was overcast and the second was sunny. We found that when direct bright sunlight shone on the cuvette, we could see the microbubbles and BAM complexes with the flashlight on or off. There was no significant change in image quality or ability to capture BAM complexes, but we observed more convection of free microbubbles in the cuvette during the magnetic capture step. To eliminate the risk that this might affect the results, we turned the vehicle around to shield the cuvette from bright sunlight before recording. In the future, we plan to add a light shield to the apparatus.

## Supporting information

Supplementary Excel Sheets

## Figure creation

All figures are original and were created using the following software: PowerPoint (Figures 1, 2, and 8); Excel (Supplementary Figure 6 and Supplementary Tables 1-13), MATLAB (Figures 3 C,D; Figure 5C and Supplementary Figures 3 and 7), and Origin (Figures 3A,B,E,F; 5A,B, 6, 7C,D, and Supplementary Figures 2, 4, and 5). ImageJ for track movement length analysis in Figure 3. Biorender was used for Supplementary Figure 8.

## Code availability

The MATLAB code used for this study is available in the Supplementary Information. The software utilized is MATLAB R2023a.

## Supplementary Information

Supplementary Information includes a PDF document with Supplementary Figures and Tables, MATLAB scripts for video analysis, Additional files include an Excel workbook with datasests for numerical figures and tables.

## Author contributions

The authors had the following CRediT taxonomy contributor roles: CW: Investigation, methodology, software, visualization, writing - original draft, and writing - review & editing; ES: investigation, validation, writing – review & editing; NE: visualization, writing – review & editing; JC: investigation, validation; WW: investigation, validation; PM: methodology, investigation, funding acquisition, writing – review & editing; DD: methodology, investigation, funding acquisition, resources, validation, writing – review & editing; JNA: conceptualization, resources, formal analysis, methodology, project administration, software, supervision, visualization, funding acquisition, writing - original draft, and writing – review & editing.

## Data Availability Statement

All data generated or analyzed during this study are included in this published article and its supplementary information files. A movie of a paramedic performing the test is available from the corresponding author upon reasonable request.

## Acknowledgements

This research was financially supported by “Deploying Buoyant-Analyte-Magnetic (BAM) Assays for Ultrasensitive Yet Rapid Detection at the Point-of-Care (POC),” COVID Testing Research Seed Grant Program from Clemson University and “Rising to the Separations and Diagnostic Challenge with Buoyancy and Magnetism (BAM),” Agilent Technologies Inc., Applications and Core Technology – University Relations (ACT-UR) Project 4599, “STI BAD: Sexually Transmitted Infection Biobank for Advanced Diagnostics,” Prisma Health Seed Grant, and the Clemson University Creative Inquiry and Summer UR programs Project 1852. We also thank Dr. Carolina Livi, PhD. at Akadeum Life Sciences and Southern Oregon University for the helpful discussions. We thank Dr. Wook Beltran, DO, MPH at Prisma Health Emergency Medicine for helping to set up the community paramedic study.

## References

1. Kost, G. J. The Impact of Increasing Disease Prevalence, False Omissions, and Diagnostic Uncertainty on Coronavirus Disease 2019 (COVID-19) Test Performance. 10.5858/arpa.2020-0716-SA (2021) doi:10.5858/arpa.2020-0716-SA.

2. Winnett, A. et al. SARS-CoV-2 Viral Load in Saliva Rises Gradually and to Moderate Levels in Some Humans. Preprint at 10.1101/2020.12.09.20239467 (2020).

3. Savela, E. S. et al. Quantitative SARS-CoV-2 Viral-Load Curves in Paired Saliva Samples and Nasal Swabs Inform Appropriate Respiratory Sampling Site and Analytical Test Sensitivity Required for Earliest Viral Detection. J. Clin. Microbiol. 60, e01785–21 (2022).

4. Kahn, M. et al. Performance of antigen testing for diagnosis of COVID-19: a direct comparison of a lateral flow device to nucleic acid amplification based tests. BMC Infect. Dis. 21, 798 (2021).

5. Target product profiles for priority diagnostics to support response to the COVID-19 pandemic v. 1.0. 2020. (2021).

6. Krenn, F. et al. Ten rapid antigen tests for SARS-CoV-2 widely differ in their ability to detect Omicron-BA.4 and -BA.5. Med. Microbiol. Immunol. (Berl.) 212, 323–337 (2023).

7. Ferretti, L. et al. Digital measurement of SARS-CoV-2 transmission risk from 7 million contacts. Nature 626, 145–150 (2024).

8. DeWolf, S. et al. SARS-CoV-2 in immunocompromised individuals. Immunity 55, 1779–1798 (2022).

9. McGarry, B. E., Gandhi, A. D. & Barnett, M. L. Covid-19 Surveillance Testing and Resident Outcomes in Nursing Homes. N. Engl. J. Med. 388, 1101–1110 (2023).

10. McGarry, B. E., SteelFisher, G. K., Grabowski, D. C. & Barnett, M. L. COVID-19 Test Result Turnaround Time for Residents and Staff in US Nursing Homes. JAMA Intern. Med. 181, 556–559 (2021).

11. Food, U. S. & Administration, D. Simoa SARS-CoV-2 N Protein Antigen Test. US Food Drug Adm. Silver Spring MD USA (2021).

12. Shan, D. et al. N-protein presents early in blood, dried blood and saliva during asymptomatic and symptomatic SARS-CoV-2 infection. Nat. Commun. 12, 1931 (2021).

13. Sarah E. Smith-Jeffcoat, M. P. H., et al. SARS-CoV-2 Viral Shedding and Rapid Antigen Test Performance — Respiratory Virus Transmission Network, November 2022–May 2023. MMWR Morb. Mortal. Wkly. Rep. 73, (2024).

14. Pumford, E. A. et al. Developments in integrating nucleic acid isothermal amplification and detection systems for point-of-care diagnostics. Biosens. Bioelectron. 170, 112674 (2020).

15. Varlamov, D. A., Blagodatskikh, K. A., Smirnova, E. V., Kramarov, V. M. & Ignatov, K. B. Combinations of PCR and Isothermal Amplification Techniques Are Suitable for Fast and Sensitive Detection of SARS-CoV-2 Viral RNA. Front. Bioeng. Biotechnol. 8, 604793 (2020).

16. Wang, C., Satterfield, E., Livi, C. B., Dean, D. & Anker, J. N. Rising to the ultrasensitive rapid diagnostic challenge with buoyant-analyte-magnetic (BAM) assays. Npj Biosensing 2, 34 (2025).

17. Lu, S. et al. The SARS-CoV-2 nucleocapsid phosphoprotein forms mutually exclusive condensates with RNA and the membrane-associated M protein. Nat. Commun. 12, 502 (2021).

18. Ham, R. E. et al. Efficient SARS-CoV-2 Quantitative Reverse Transcriptase PCR Saliva Diagnostic Strategy utilizing Open-Source Pipetting Robots. JoVE J. Vis. Exp. e63395 (2022) doi:10.3791/63395.

19. Olsen, D. Aa., et al. Quantifying SARS-CoV-2 nucleocapsid antigen in oropharyngeal swabs using single molecule array technology. Sci. Rep. 11, 20323 (2021).

20. Ren, A. et al. Ultrasensitive assay for saliva-based SARS-CoV-2 antigen detection. Clin. Chem. Lab. Med. CCLM 60, 771–777 (2022).

21. Groh, A. M. et al. Kinetics of SARS-CoV-2 infection biomarkers in a household transmission study. Sci. Rep. 14, 12365 (2024).

22. Jain, P. K., Lee, K. S., El-Sayed, I. H. & El-Sayed, M. A. Calculated Absorption and Scattering Properties of Gold Nanoparticles of Different Size, Shape, and Composition: Applications in Biological Imaging and Biomedicine. J. Phys. Chem. B 110, 7238–7248 (2006).

23. Khlebtsov, B. N., Tumskiy, R. S., Burov, A. M., Pylaev, T. E. & Khlebtsov, N. G. Quantifying the Numbers of Gold Nanoparticles in the Test Zone of Lateral Flow Immunoassay Strips. ACS Appl. Nano Mater. 2, 5020–5028 (2019).

24. Zhan, L. et al. The Role of Nanoparticle Design in Determining Analytical Performance of Lateral Flow Immunoassays. Nano Lett. 17, 7207–7212 (2017).

25. Cho, D. G. et al. High-Speed Lateral Flow Strategy for a Fast Biosensing with an Improved Selectivity and Binding Affinity. Sensors 18, 1507 (2018).

26. Pierres, A., Benoliel, A.-M. & Bongrand, P. Measuring the Lifetime of Bonds Made between Surface-linked Molecules (∗). J. Biol. Chem. 270, 26586–26592 (1995).

27. Choi, J. R. et al. Polydimethylsiloxane-Paper Hybrid Lateral Flow Assay for Highly Sensitive Point-of-Care Nucleic Acid Testing. Anal. Chem. 88, 6254–6264 (2016).

28. Dong, R., Yi, N. & Jiang, D. Advances in single molecule arrays (SIMOA) for ultra-sensitive detection of biomolecules. Talanta 270, 125529 (2024).

29. The development of an artificial saliva for in vitro amalgam corrosion studies - DARVELL - 1978 - Journal of Oral Rehabilitation - Wiley Online Library. https://onlinelibrary.wiley.com/doi/abs/10.1111/j.1365-2842.1978.tb00390.x.

30. Frontiers | Repurposing a SARS-CoV-2 surveillance program for infectious respiratory diseases in a university setting. https://www.frontiersin.org/journals/public-health/articles/10.3389/fpubh.2023.1168551/full.

31. Ham, R. E. et al. Identifying SARS-CoV-2 Variants of Concern through Saliva-Based RT-qPCR by Targeting Recurrent Mutation Sites. Microbiol. Spectr. 10, e00797–22 (2022).

