## Supplementary Information for "Developing Buoyant-Analyte-Magnetic (BAM) Assays for Ultrasensitive Yet Rapid Point-of-Care Detection"

**Contents:	Page**

**Figures:**

Supplementary Figure 1: Separation and enrichment protocol for larger buoyant microbubbles.	2

Supplementary Figure 2: Calibration curves for real PCR-negative saliva spiked with N-protein with manual and semi-automated counting.	3

Supplementary Figure 3: Track analysis for simplified BAM POC assays using cotton swab.	4

Supplementary Figure 4: Box plot for patient saliva using larger POC assay with handmade cotton swab filter.	5

Supplementary Figure 5: Linear regression to pooled data from four independent tests.	6

Supplementary Figure 6: Time-Dependent N-protein Signal Decay in Saliva.	7

Supplementary Figure 7: Images of BAM complexes recorded every 5 seconds for 20 seconds following the key frame during three independent 10 fg/mL concentration tests.	8

Supplementary Figure 8. BAM assay instruction booklet.	9

Supplementary Figure 9: Photos of workflow steps in the Iteration 2 protocol	10

**Tables:**

Supplementary Table 1: Dataset for Figure 3E.	11

Supplementary Table 2: Dataset for Figure 3F.	11

Supplementary Table 3: Costs of device and disposables.	11

Supplementary Table 4: Dataset for Supplementary Figure 2.	12

Supplementary Table 5: Dataset for Supplementary Figure 4.	12

Supplementary Table 7: Dataset for Figure 5A.	13

Supplementary Table 7: Dataset for Figure 5B.	13

Supplementary Table 8: Dataset for Figure 6A.	14

Supplementary Table 9: Dataset for Figures 6B and 7D.	14

Supplementary Table 10: BAM assay results of serially diluted false negative samples.	14

Supplementary Table 11: Dataset for Figure 7C and Supplementary Figure 5.	15

Supplementary Table 12: Dataset for Supplementary Figure 6.	16

Supplementary Table 13: Dataset for Supplementary Figure 7.	16

**Matlab Scripts:**	17-24

**Supplementary Figure 1: Separation and enrichment protocol for larger buoyant microbubbles. A)** Initial 10 mL syringe setup, showing the buoyant microbubbles solution after the initial 30-minute flotation towards the tip. **B)** Differential fractionation after inverting the syringe and allowing 5 minutes of static separation. **C)** Final target larger buoyant microbubble concentrated near the plunger end, before expelling the solution containing smaller buoyant microbubble from the tip.

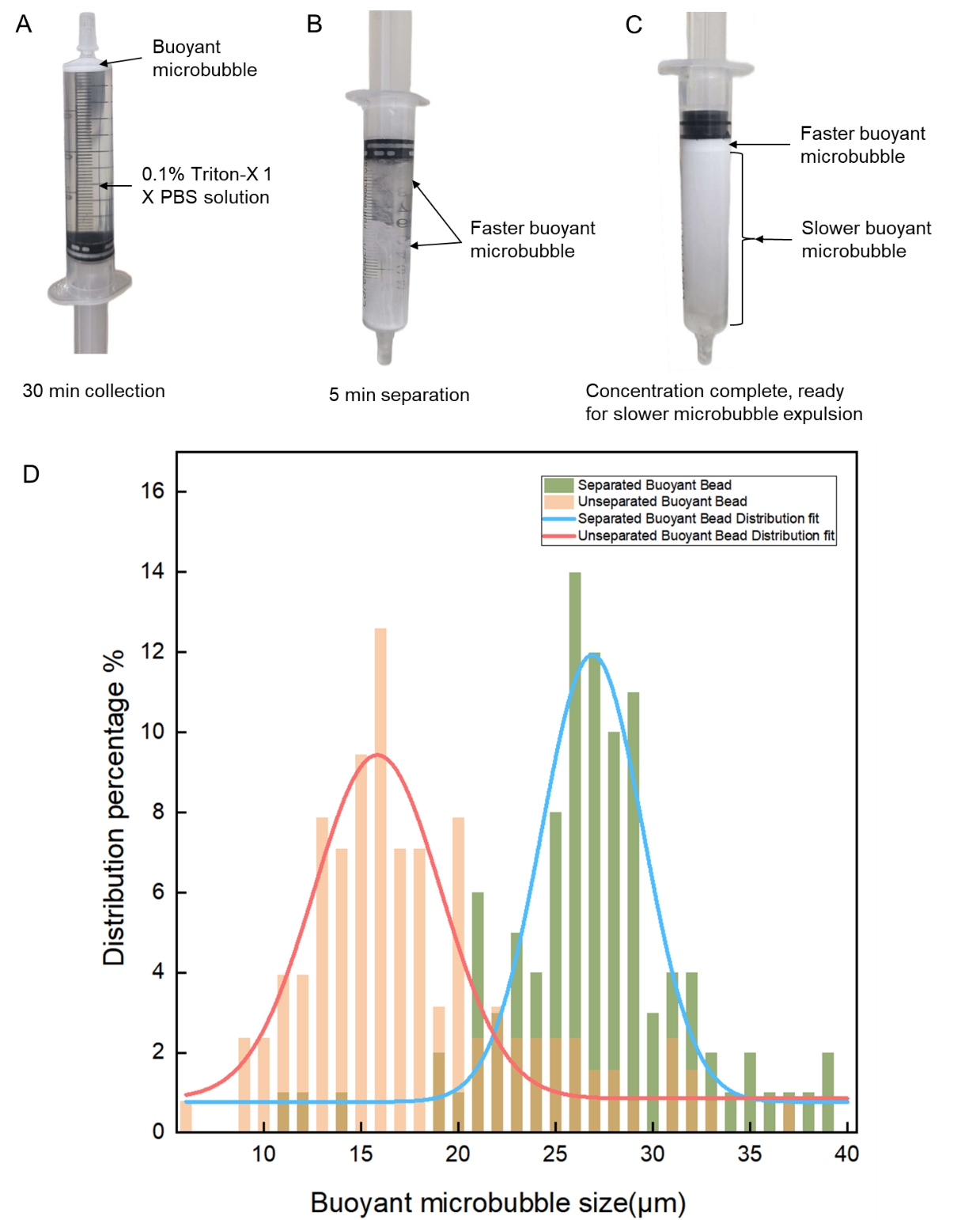

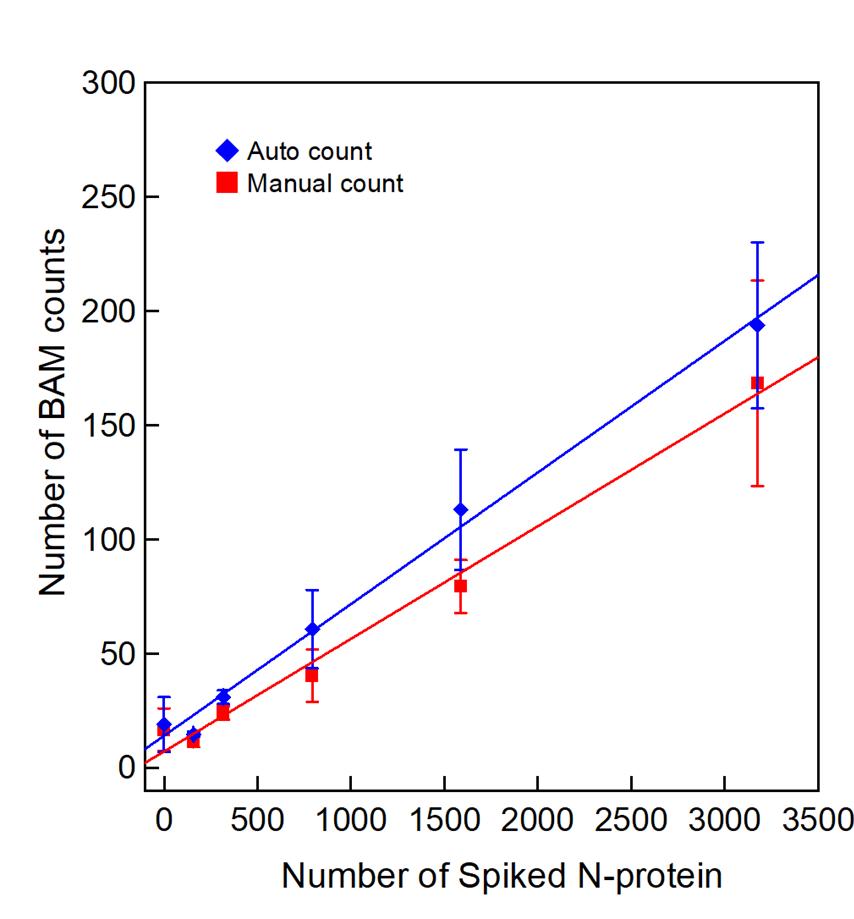

**Supplementary Figure 2:** Calibration curve for real PCR-negative saliva spiked with N-protein at 0, 0.5 fg/mL, 1 fg/mL, 2.5 fg/mL, 5 fg/mL, 10 fg/mL. The auto counts showed 5.8% capture rate, the manual count showed 4.9% capture rate. Data in Supplementary Table 4.

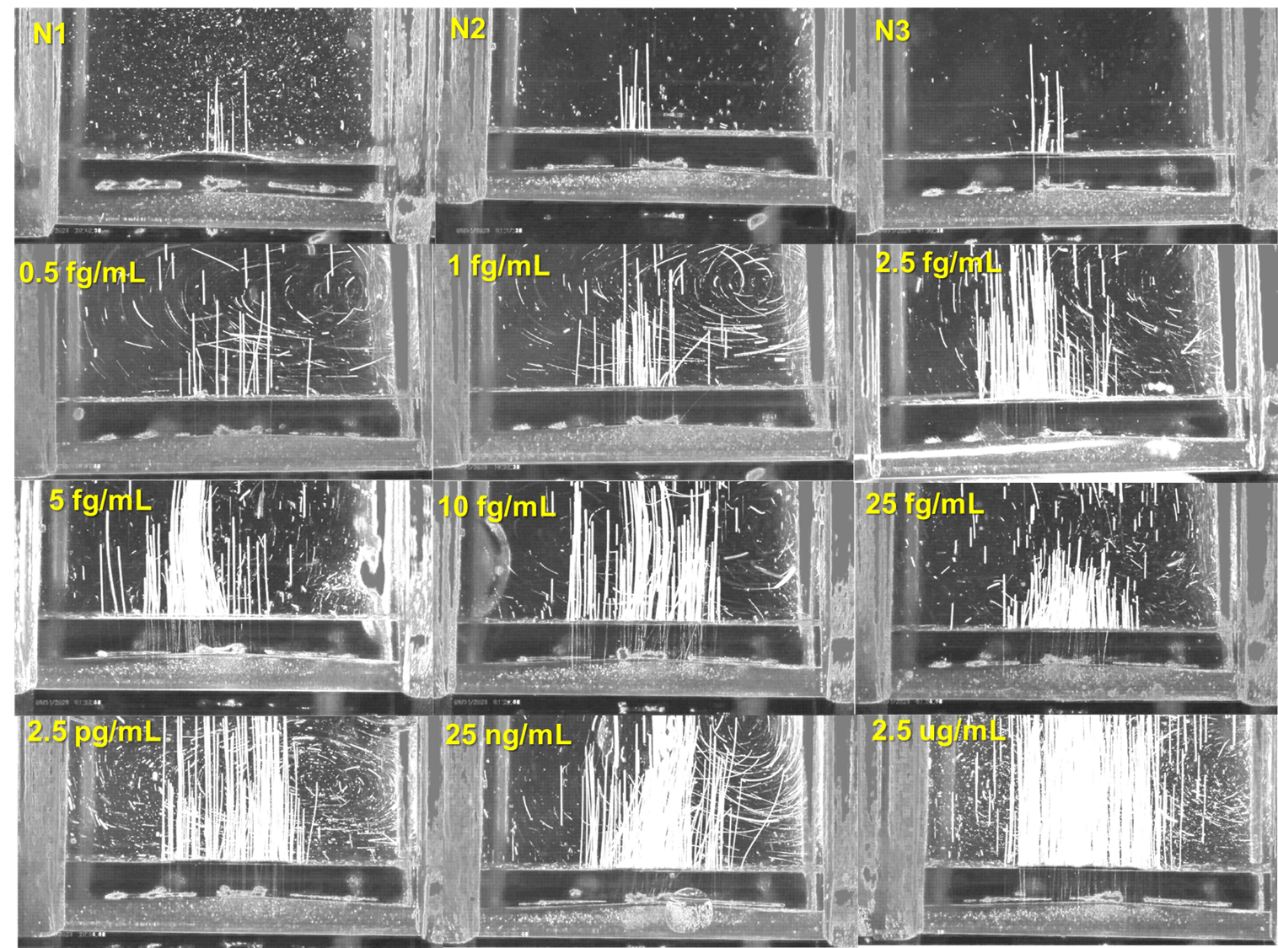

**Supplementary Figure 3:** Track analysis for simplified BAM POC in PCR-negative patient saliva assays using a cotton swab instead of a 10 µm filter (0 fg/mL to 2.5 µg/mL N-protein).

**Supplementary Figure 4:** Box plot for patient saliva sample test, with handmade cotton swab filter. Negative mean 12.8, standard deviation 19.3. Positive mean 106.5, standard deviation 52.2, indeterminate mean 40.3, standard deviation 22.0. Dataset in Supplementary Table 5.

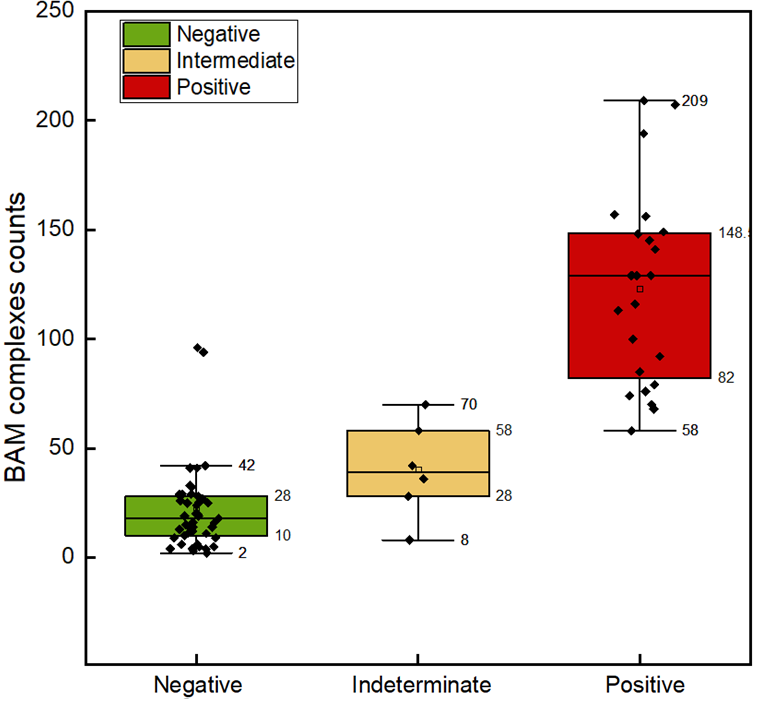

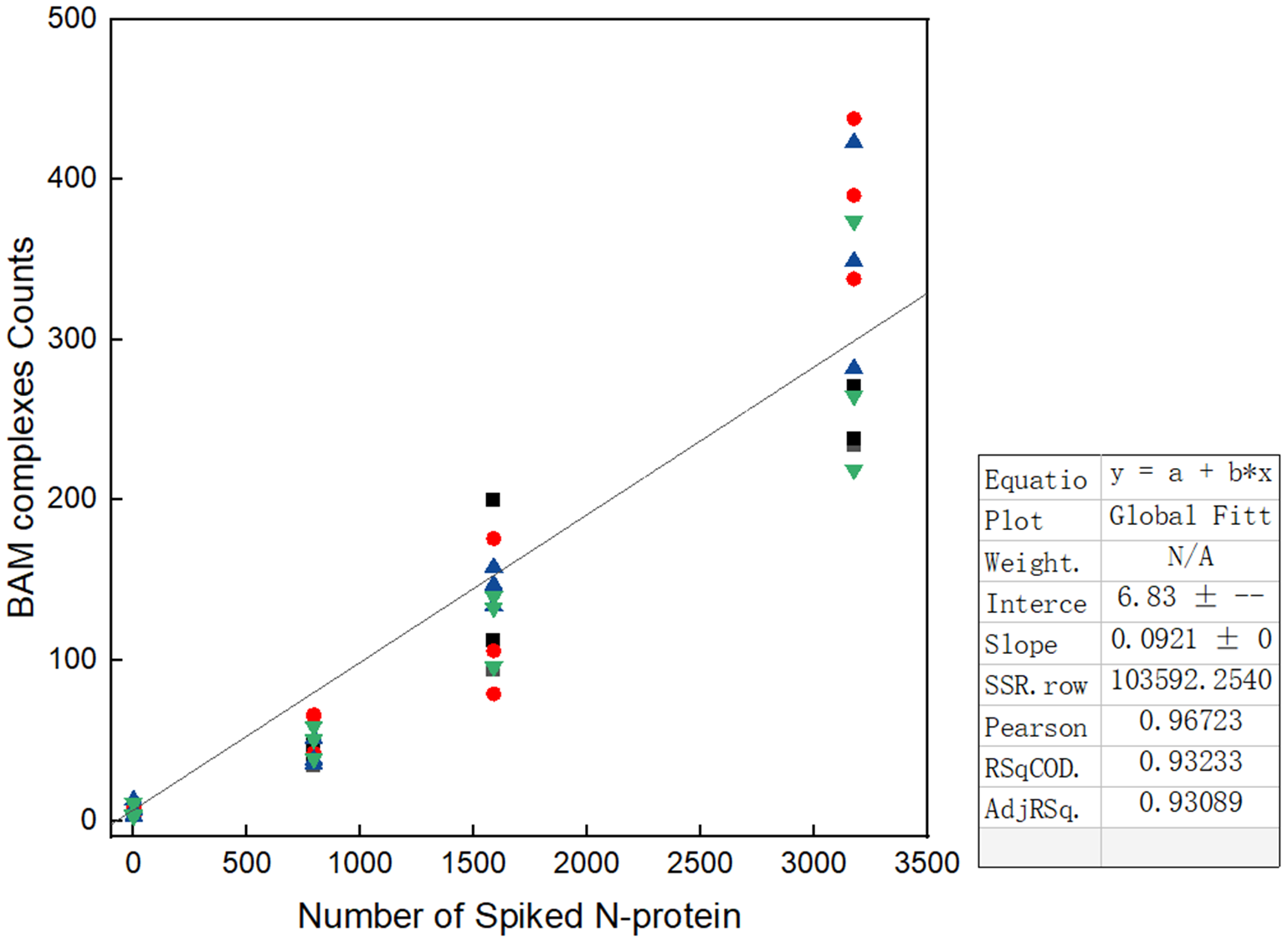

**Supplementary Figure 5:** Linear regression to pooled data from four independent tests conducted by different operators across varied experimental environments (Lab Researcher, Emergency Department technician in lab, Emergency Department technician in hospital, and community paramedic in van. The curves were generated using mock saliva spiked with 0, 2.5, 5, and 10 fg/mL N-protein. Dataset in Supplementary Table 11.

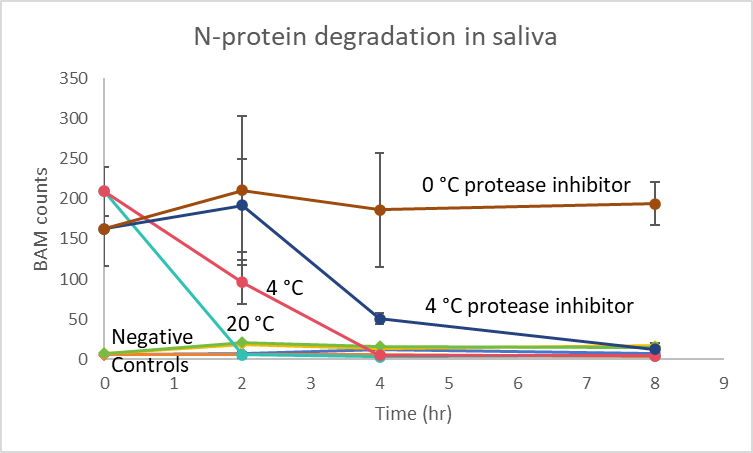

**Supplementary Figure 6: Time-Dependent N-protein Signal Decay in Saliva.** The graph illustrates the BAM complexes signal stability in 10 fg/mL N-protein over 8 hours as influenced by temperature 0 °C, 4 °C, 20 °C and the presence of a protease inhibitor. Dataset in Supplementary Table 12.

**Supplementary Figure 7:** **Images of BAM complexes recorded every 5 seconds for 20 seconds following the key frame** during three independent 10 fg/mL concentration tests. These images were used for subsequent manual counting and trajectory analysis. A-D correspond to the images of complexes captured at t=5 s, 10 s, 15 s, and 20 s during the first test run. E-H shows the results from the second test, and I-N represents the results from the third test. BAM count data shown in Supplementary Table 13.

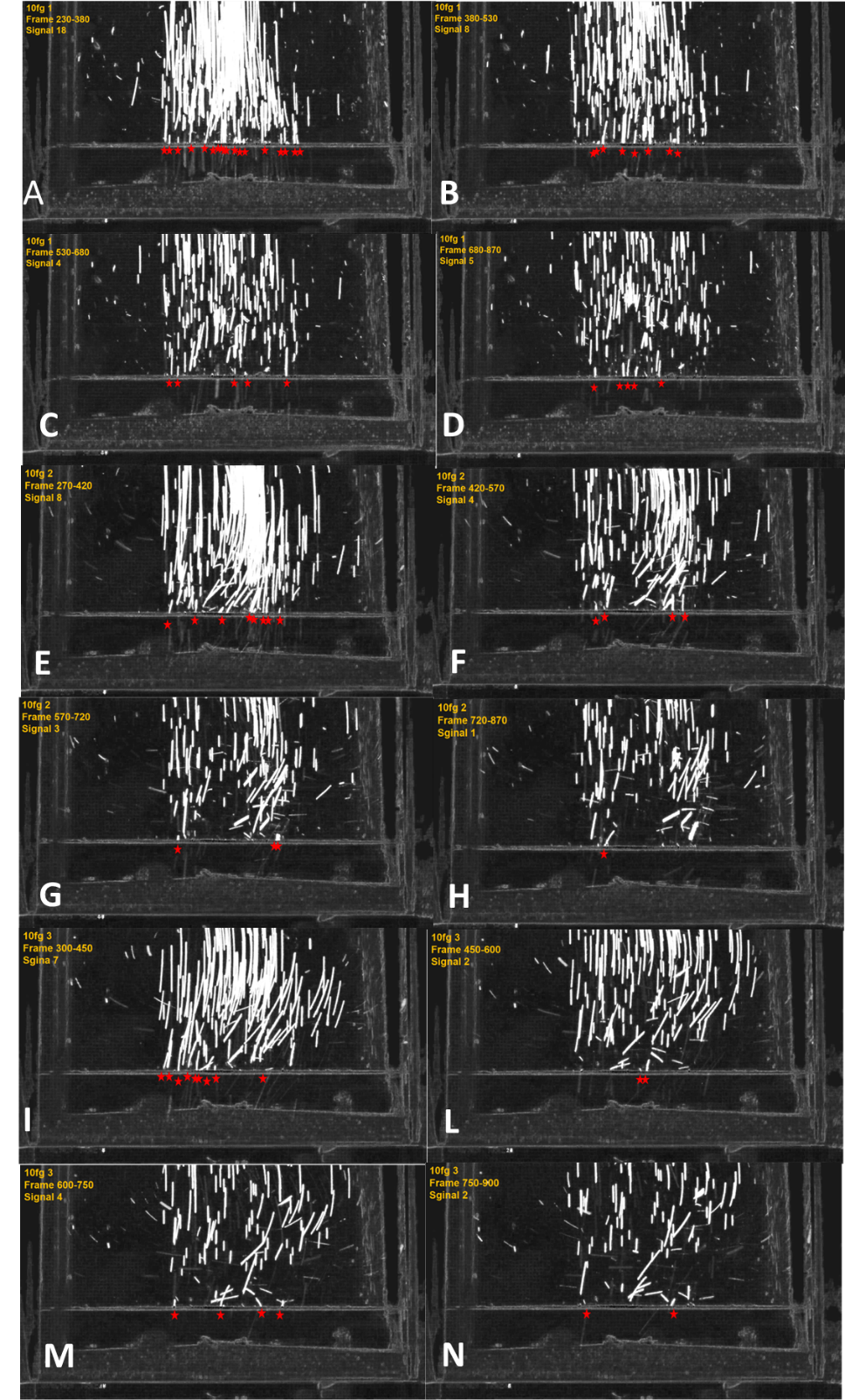

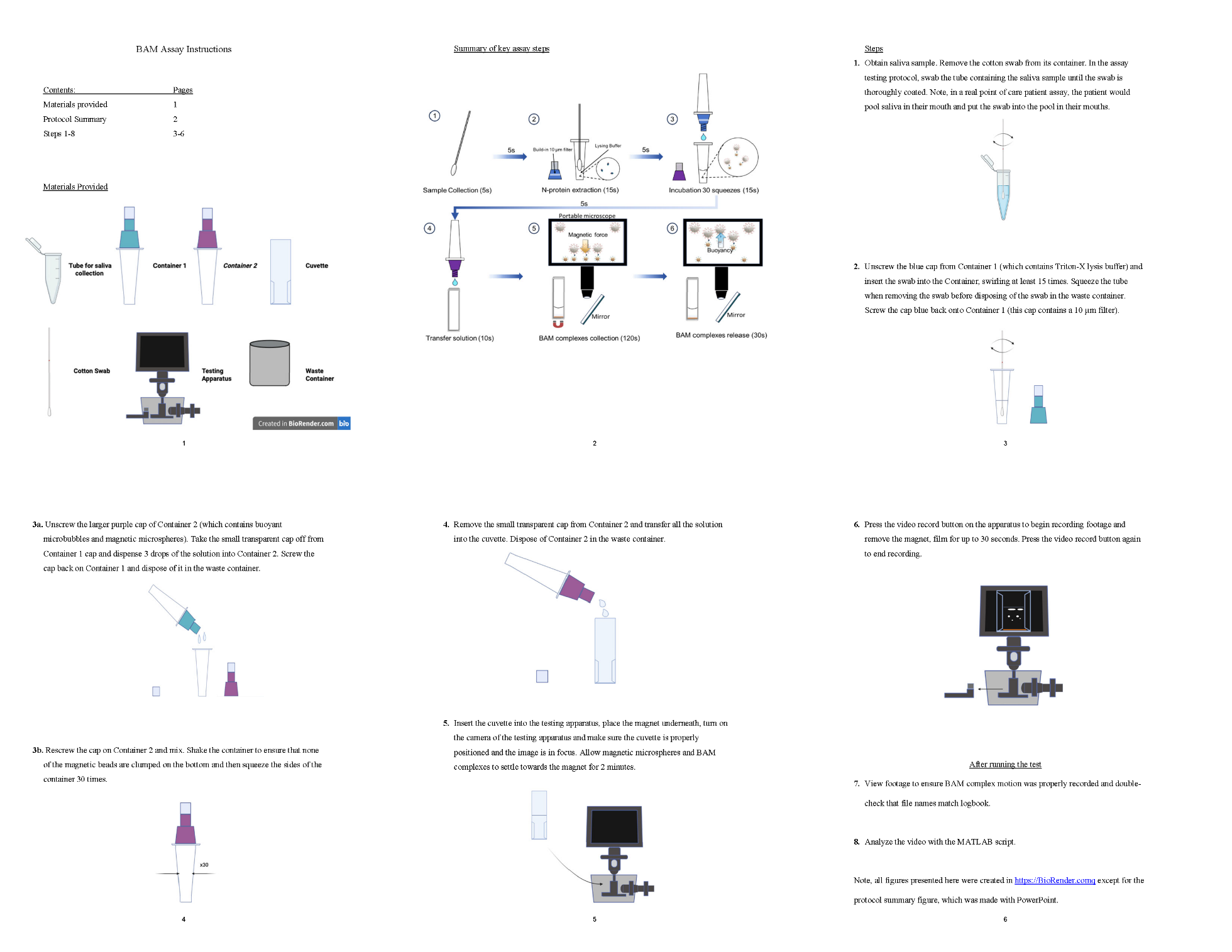

**Supplementary Figure 8.** BAM assay instruction booklet.

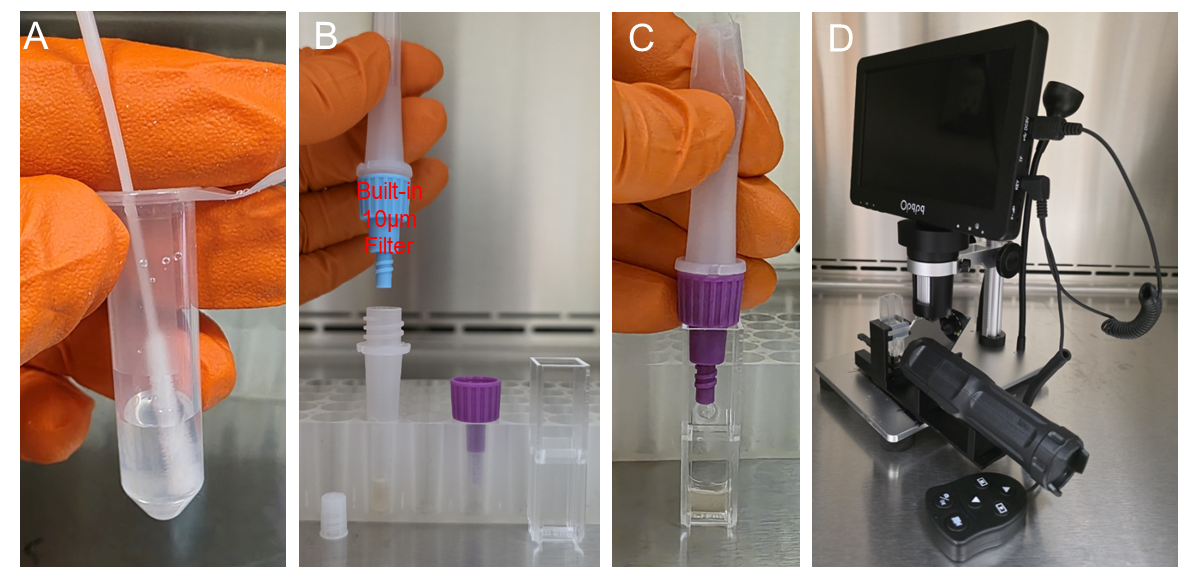

**Supplementary Figure 9:** **Photos of workflow steps in the Iteration 2 protocol**. **A)** A saliva swab was placed into a lysis/inactivation buffer, squeezed to extract saliva, and capped with a blue filter cap (10 µm filter). **B)** 3 drops (131 ± 9 µL) of the lysed specimen added through filter cap to buoyant and magnetic beads + Squeezed 30 times. **C)** the mixture was poured into cuvette, placed in stand, and the microscope adjusted to ensure proper alignment. **D)** The cuvette was left for 2 minutes for free microbubbles to rise and solution to clarify, and then the magnet was removed and rising BAM complexes counted.

**Supplementary Table 1:** Dataset for Figure 3E.

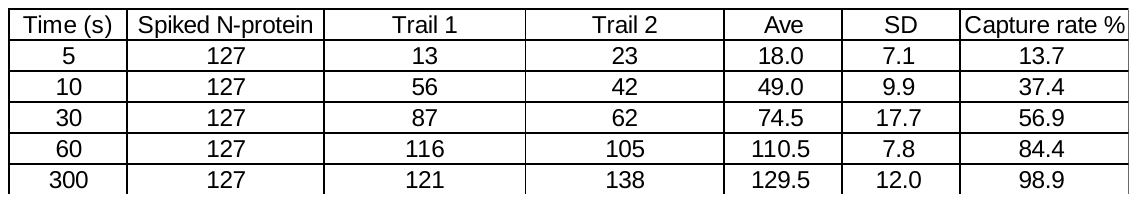

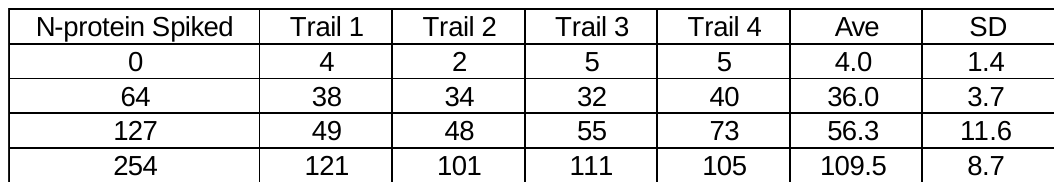

**Supplementary Table 2:** Dataset for Figure 3F.

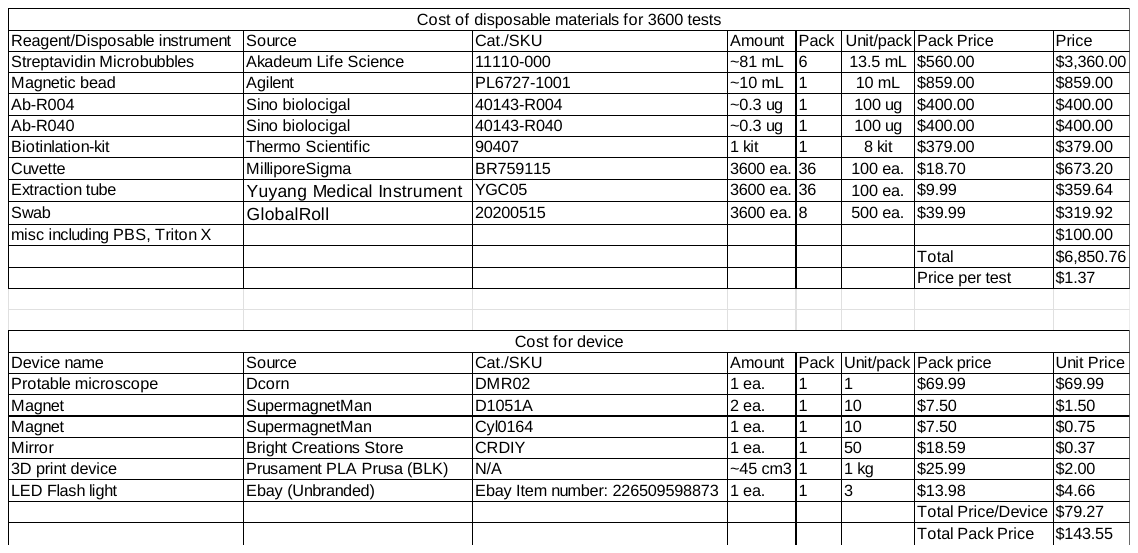

**Supplementary Table 3:** Cost of device and disposables.

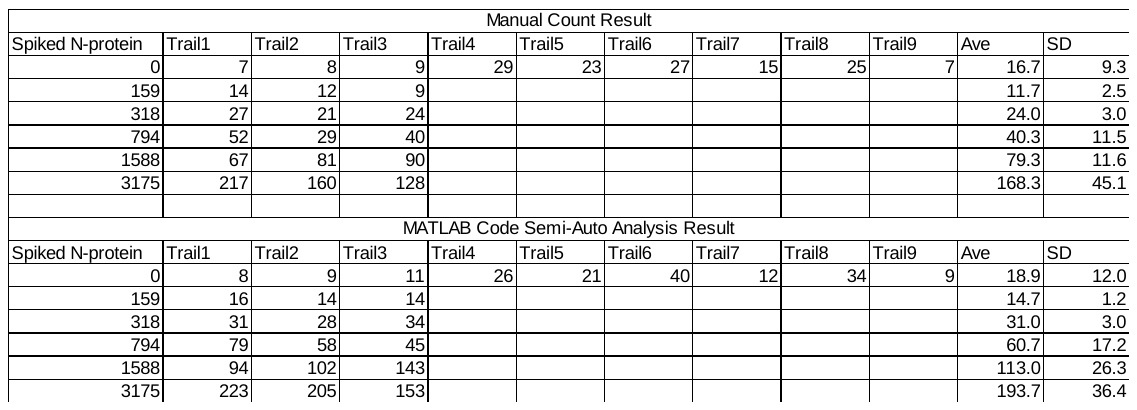

**Supplementary Table 4:** Dataset for Supplementary Figure 2 calibration curve. Note, Trials 1-3 were freshly prepared negative saliva spiked with N-protein to 0, 0.5, 1, 2.5, 5, and 10 fg/mL. Trials 4-6 and 7-9 were from two frozen PCR-negative saliva specimens.

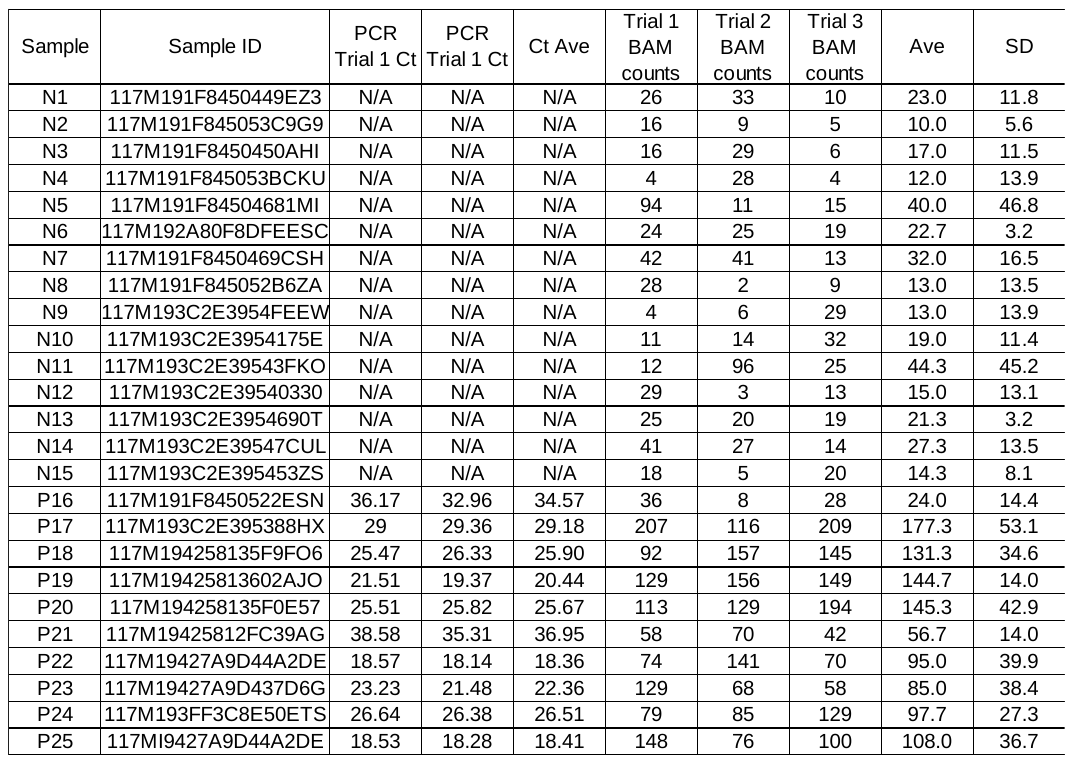

**Supplementary Table 5:** Dataset for Supplementary Figure 4, "Iteration 1.5," using the instrumentation and protocols from iteration 2, except using the Salivatte filter from Iteration 1 in place of the 10 µm filters.

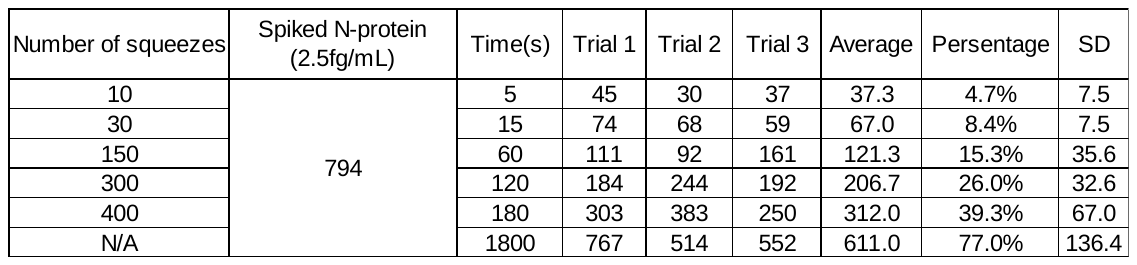

**Supplementary Table 6:** Dataset for Figure 5A.

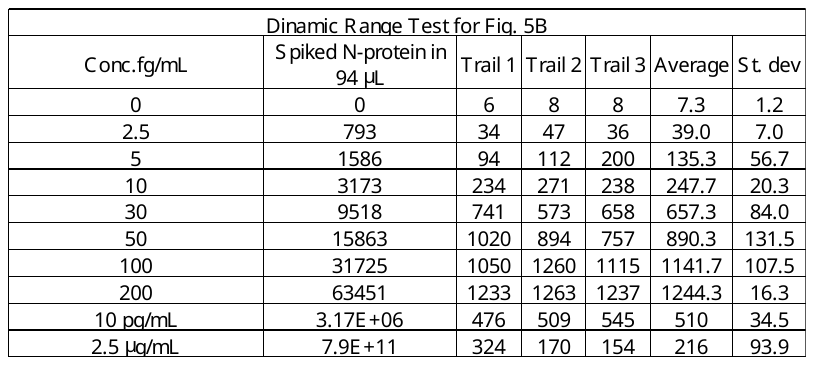

**Supplementary Table 7:** Dataset for Figure 5B.

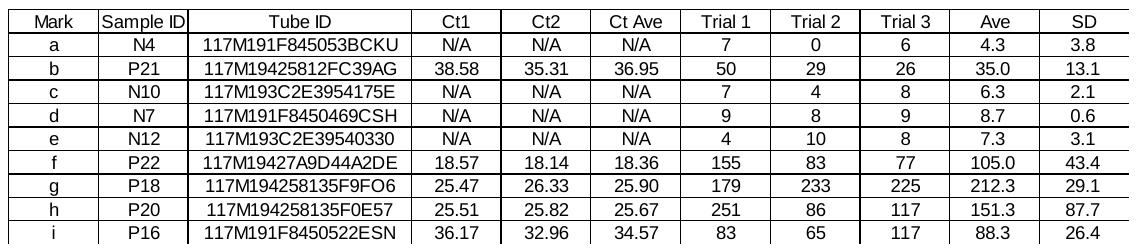

**Supplementary Table 8:** Dataset for Figure 6A.

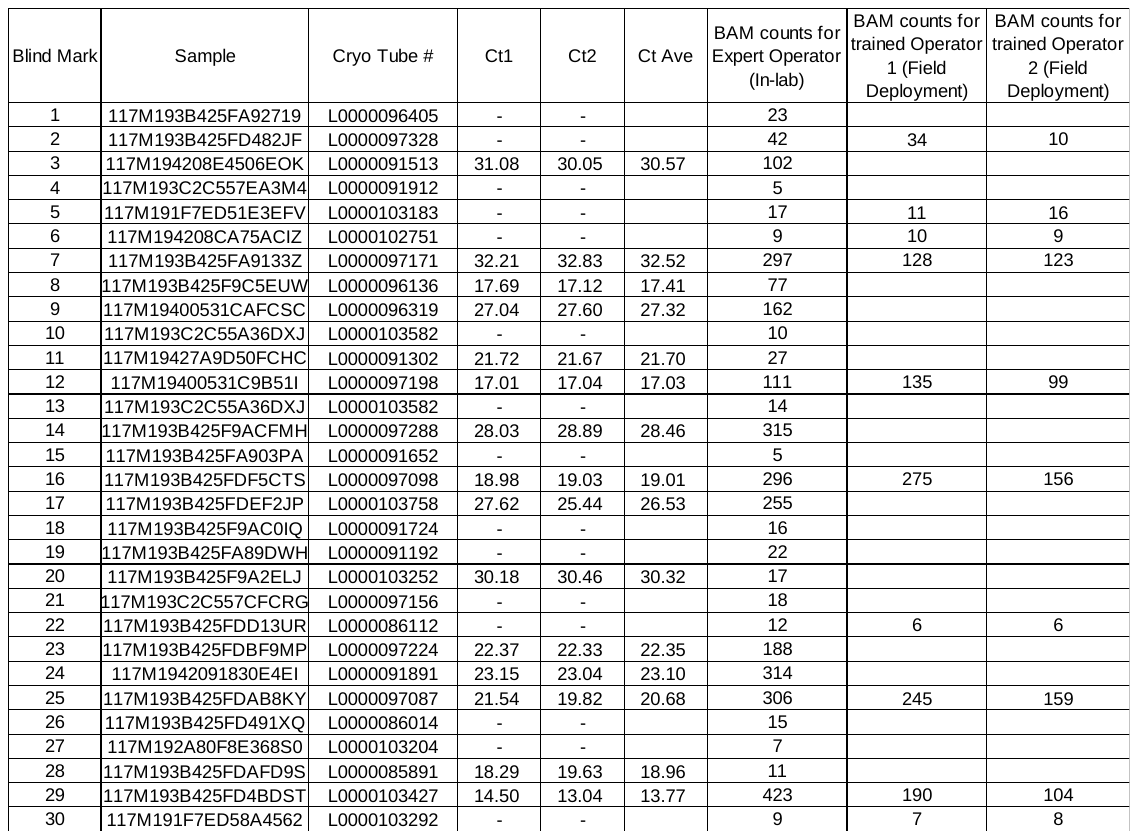

**Supplementary Table 9:** Dataset for Figures 6B and 7D.

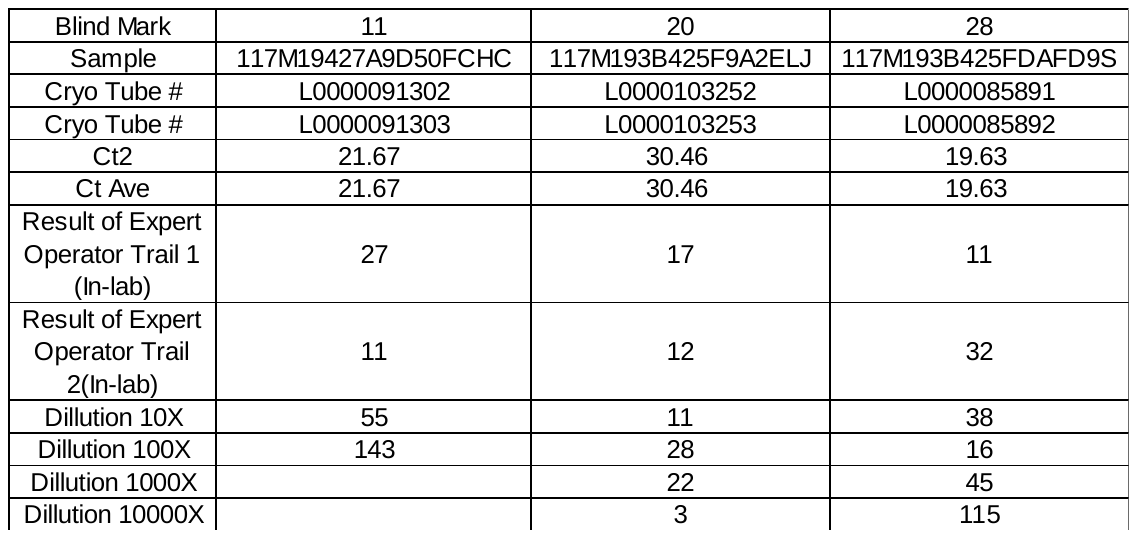

**Supplementary Table 10:** BAM assay results of serially diluted false negative samples.

**Supplementary Table 11:** Dataset for Figure 7C and Supplementary Figure 5. Pooled 0 concentration value is 6.8 ±3.2. Note that the first “in-lab” data is the same as for in-lab calibration Fig. 5B and Supplementary Table 6.

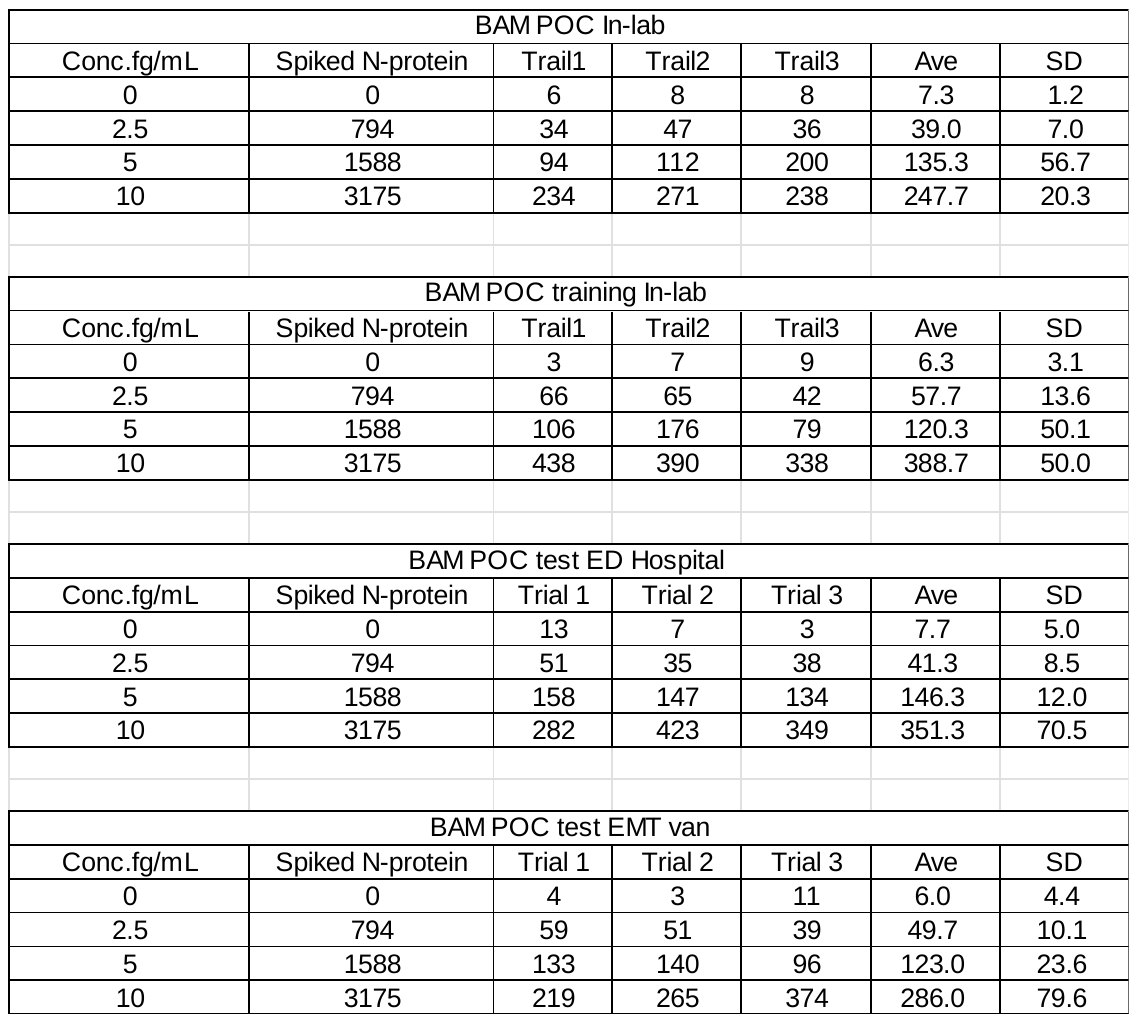

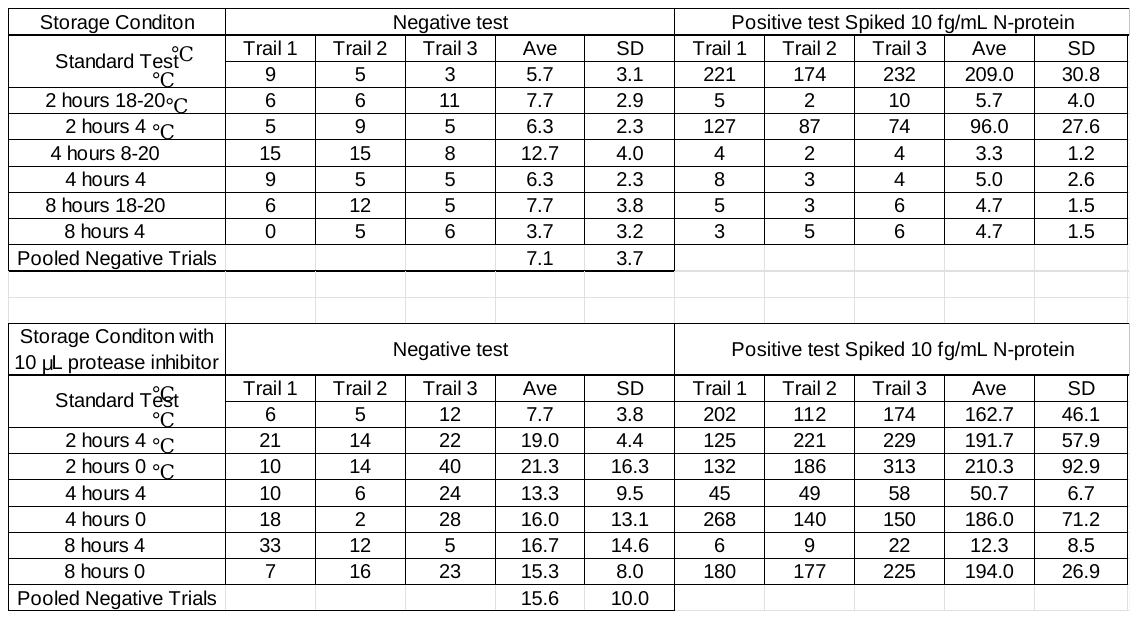

**Supplementary Table 12:** Dataset for Supplementary Figure 6.

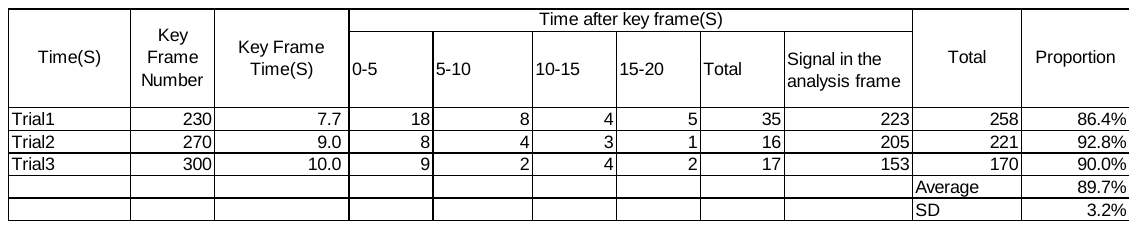

**Supplementary Table 13.** Dataset for Supplementary Figure 7.

**Matlab scripts.**

Below are three Scripts, “Video_particle_ct,” “Vid_minI,” and “Charlie_count”

Each script can be saved as a separate .m file and run in Matlab.

“Video_particle_ct” loads the video and calls the other two functions.

“Vid_minI” analyzes the video, selects a key frame, and subtracts off the background (defined for each pixel as the minimum intensity for that pixel in the movie).

“Charlie_count” counts the number of particles in the background-subtracted frame, with user input if needed for manually removing non-particles or highlighting particles that are missed.

**function [MinI, Im_ct]=Video_particle_ct();**

close all;

playerHandles = findall(0, 'Type', 'figure');

for i = 1:length(playerHandles)

if strcmp(get(playerHandles(i), 'Name'), 'Movie')

close(playerHandles(i)); % Close all

end

end

[Fname2, pth]=uigetfile('*.*');

cd(pth);

implay(Fname2)

s_frame= input('Enter Start frame # ');

e_frame=input('Enter End frame # ');

[NumFrames,Fname, Io01, AveI, MinI, MaxI,V1,Io01min]=Vid_minI(s_frame,e_frame,Fname2);

ct_frame=input('Enter count frame #');

Im_ct=read(V1,ct_frame);

figure; imshow(Im_ct);

Im_ct_r(:,:)=Im_ct(:,:,1);

Charlie_count(Im_ct_r-Io01min);

**function [NumFrames,Fname, Io01, AveI, MinI, MaxI,V1,Io01min]=Vid_minI(s_frame, e_frame, Fname)**

if nargin<1,

s_frame=1; %start frame

end;

V1=VideoReader(Fname);

NumFrames = get(V1,'NumberOfFrames')

if nargin<2,

e_frame=NumFrames; %end frame

end;

I=read(V1,s_frame);

f1=figure();

imshow(I);

Io01f1=read(V1,s_frame);

AveI(1)=mean(mean(Io01f1(:,:,1)));

MinI(1)=min(min(Io01f1(:,:,1)));

MaxI(1)=max(max(Io01f1(:,:,1)));

Io01min(:,:)=Io01f1(:,:,1); %min red

Io01max(:,:)=Io01f1(:,:,1); %max red

for i=s_frame+1:e_frame %frame number

Io01=read(V1,i);

AveI(i+1-s_frame)=mean(mean(Io01(:,:,1)));

MinI(i+1-s_frame)=min(min(Io01(:,:,1)));

MaxI(i+1-s_frame)=max(max(Io01(:,:,1)));

Io01min(:,:)=min(Io01min,Io01(:,:,1));

Io01max(:,:)=max(Io01max,Io01(:,:,1));

end

figure; plot(s_frame:e_frame,AveI); hold; plot(s_frame:e_frame,MinI); plot(s_frame:e_frame,MaxI);

xlabel('frame number');

ylabel('Intensity');

figure; imshow(Io01min);

figure; imshow(Io01max-Io01min);

function []=Charlie_count(originalImage, minbgimage)

% This command counts particles (e.g., BAM complexes) in an image with some user input.

% Usually the image is selected in “Video_particle_ct()” which selects a key frame in a

% movie and subtracts the background (defined for each pixel as the minimum intensity

% the pixels exhibits in time in the video clip.

% Step 1: Input an image if not already entered

if nargin < 1

[filename, pathname] = uigetfile({'*.jpg;*.png;*.bmp', 'Image Files (*.jpg, *.png, *.bmp)'});

if isequal(filename, 0)

disp('User canceled the file selection.');

return;

end

fullpath = fullfile(pathname, filename);

disp(['Selected file: ', fullpath]); % Debug: Display selected file path

% Step 2: Read and display the image

originalImage = imread(fullpath);

end

figure;

imshow(originalImage);

title('Original Image');

% Loop for parameter setting

while true

% Step 3: Convert to grayscale

grayImage = im2gray(originalImage);

% Gray map

figure;

imshow(grayImage);

title('Grayscale Image');

% Step 4: Crop Screenshot

rect = getrect; % Select region

selectedRegion = imcrop(grayImage, rect); % Crop

% Show crop region

figure;

imshow(selectedRegion);

title('Selected Region');

% Step 5: Thresholding to create a binary image

signalStrength = input('Enter the threshold for signal intensity (0-255): ');

binaryImage = selectedRegion > signalStrength; % Binary the cropregion

% Debug: Check binary image

disp(['Binary image class: ', class(binaryImage)]); % Check data type

disp(['Binary image size: ', num2str(size(binaryImage))]); % Check size

disp(['Sum of binaryImage: ', num2str(sum(binaryImage(:)))]); % Check number of detected signals

% Optional: Filter by area (adjust the area threshold as needed)

minArea = input('Enter the minimum area of the signal in pixels: ');

if sum(binaryImage(:)) == 0

disp('No signals detected. Try adjusting the threshold value or area.');

continue; % Go back to parameter setting

end

% Use bwconncomp instead of bwlabel

cc = bwconncomp(binaryImage);

stats = regionprops(cc, 'Area', 'BoundingBox');

% Step 6: Mark and count circles

count = 0;

outputImage = selectedRegion; % Output

for i = 1:length(stats)

if stats(i).Area >= minArea

count = count + 1;

bbox = stats(i).BoundingBox;

centerX = bbox(1) + bbox(3) / 2;

centerY = bbox(2) + bbox(4) / 2;

radius = bbox(3) / 2;

outputImage = insertShape(outputImage, 'Circle', ...

[centerX, centerY, radius], ...

'Color', 'red', 'LineWidth', 2);

end

end

% Step 7: Display automatic results before manual adjustment

figure;

imshow(outputImage);

title('Automatically Detected Signals (Red Circles)');

disp(['Number of automatically detected signals: ', num2str(count)]);

% Ask if the user is satisfied with the results

retry = input('Are you satisfied with the results? (y/n): ', 's');

if strcmpi(retry, 'y')

break; % Exit the loop if satisfied

end

end

% Step 7: Manual editing of results

hFig = figure;

imshow(outputImage);

title('Marked Image with Red Circles');

hold on;

% Initialize counters and storage for added points

greenCircleCount = 0; % Count of added green circles

blueArrowCount = 0; % Count of added blue arrows

greenPoints = []; % Store coordinates of green points

bluePoints = []; % Store coordinates of blue points

undoStack = []; % Stack for undo functionality

disp('Left click to add green circles, right click to add blue arrows (press Enter to finish). Press "u" to undo last action.');

while true

% Get user input for mouse clicks

[x, y, button] = ginput(1); % Get one point

if button == 1 % Left mouse button

% Add green circle

plot(x, y, 'g+', 'MarkerSize', 10, 'LineWidth', 2); % Mark point

greenCircleCount = greenCircleCount + 1; % Increment green circle count

greenPoints = [greenPoints; x, y]; % Store green point

undoStack = [undoStack; 1, x, y]; % Record action for undo

elseif button == 3 % Right mouse button

% Add blue arrow

plot(x, y, 'b+', 'MarkerSize', 10, 'LineWidth', 2); % Mark point

blueArrowCount = blueArrowCount + 1; % Increment blue arrow count

bluePoints = [bluePoints; x, y]; % Store blue point

undoStack = [undoStack; 2, x, y]; % Record action for undo

end

% Check for key press to undo or finish

if waitforbuttonpress

key = get(hFig, 'CurrentCharacter'); % Get the character of the pressed key

if strcmp(key, char(117)) % Check if 'u' key (ASCII code for 'u')

if ~isempty(undoStack)

lastAction = undoStack(end, :); % Get last action

undoStack(end, :) = []; % Remove last action from stack

% Undo the last action

if lastAction(1) == 1 % Green circle

greenCircleCount = greenCircleCount - 1; % Decrement count

% Redraw the image without the last green circle

outputImage = imshow(originalImage); hold on;

plot(greenPoints(1:end, 1), greenPoints(1:end, 2), 'g+', 'MarkerSize', 10, 'LineWidth', 2);

plot(bluePoints(:, 1), bluePoints(:, 2), 'b+', 'MarkerSize', 10, 'LineWidth', 2);

title('Marked Image with Red Circles');

hold off;

elseif lastAction(1) == 2 % Blue arrow

blueArrowCount = blueArrowCount - 1; % Decrement count

% Redraw the image without the last blue arrow

outputImage = imshow(originalImage); hold on;

plot(greenPoints(:, 1), greenPoints(:, 2), 'g+', 'MarkerSize', 10, 'LineWidth', 2);

plot(bluePoints(1:end, 1), bluePoints(1:end, 2), 'b+', 'MarkerSize', 10, 'LineWidth', 2);

title('Marked Image with Red Circles');

hold off;

end

end

elseif strcmp(key, char(13)) % Check if Enter key (ASCII code 13)

break; % Exit loop on Enter key

end

end

end

hold off;

% Step 8: Reprocess the binary image after manual modifications

cc = bwconncomp(binaryImage);

stats = regionprops(cc, 'Area', 'BoundingBox');

% Step 9: Mark and count circles again

countAfterAdjustment = 0;

outputImage = selectedRegion;

for i = 1:length(stats)

if stats(i).Area >= minArea

countAfterAdjustment = countAfterAdjustment + 1;

bbox = stats(i).BoundingBox;

centerX = bbox(1) + bbox(3) / 2;

centerY = bbox(2) + bbox(4) / 2;

radius = bbox(3) / 2;

outputImage = insertShape(outputImage, 'Circle', ...

[centerX, centerY, radius], ...

'Color', 'red', 'LineWidth', 2);

end

end

% Step 10: Draw green circles and blue arrows on the final output image

for i = 1:size(greenPoints, 1)

x = greenPoints(i, 1);

y = greenPoints(i, 2);

% Draw a green circle

outputImage = insertShape(outputImage, 'Circle', ...

[x, y, 5], ... % Small circle as a green circle

'Color', 'green', 'LineWidth', 2);

end

for i = 1:size(bluePoints, 1)

x = bluePoints(i, 1);

y = bluePoints(i, 2);

% Draw a blue arrow

outputImage = insertShape(outputImage, 'Line', [x, y, x+10, y-10], ...

'Color', 'blue', 'LineWidth', 2);

end

% Step 11: Display updated results with all circles and arrows

figure;

imshow(outputImage);

title('Final Results with Red Circles, Green Circles, and Blue Arrows');

% Step 12: Calculate total count

totalCount = count + greenCircleCount - blueArrowCount; % Total = red + green - blue

disp(['Number of automatically detected signals (red): ', num2str(count)]);

disp(['Number of manually added signals (green): ', num2str(greenCircleCount)]);

disp(['Number of blue arrows (to remove): ', num2str(blueArrowCount)]);

disp(['Total count (red + green - blue): ', num2str(totalCount)]);

% Set savepath

savePath = 'Save Path';

currentTime = datetime('now', 'Format', 'yyyyMMdd_HHmmss'); % Time

outputFilename = fullfile(savePath, [char(currentTime), '.png']); % Set time as the file name

imwrite(outputImage, outputFilename);

disp(['Final marked image saved as: ', outputFilename]);

disp('Image analysis complete.');
